# Virtual Simulated Placements in Healthcare Education: A scoping review

**DOI:** 10.1101/2023.10.12.23296932

**Authors:** Juliana Samson, Marc Gilbey, Natasha Taylor, Rosie Kneafsey

## Abstract

**Introduction:** A virtual simulated placement (VSP) is a computer-generated version of a practice placement. COVID-19 drove increased adoption of virtual technology in clinical education. Accordingly, the number of VSP publications increased from 2020. This review aims to determine the scope of this literature to inform future research questions.

**Objective:** Assess the range and types of evidence related to VSPs across the healthcare professions.

**Inclusion criteria:** Studies that focussed on healthcare students participating in VSPs. Hybrid, augmented reality (AR) and mixed reality (MR) placements were excluded.

**Methods:** Fourteen databases were searched, limited to English, and dated from 1^st^ January 2020. Supplementary searches were employed, and an updated search was conducted on 9^th^ July 2023. Themes were synthesised using the PAGER framework to highlight patterns, advances, gaps, evidence for practice and research recommendations.

**Results:** Twenty-eight papers were reviewed. All VSPs were designed in response to pandemic restrictions. Students were primarily from medicine and nursing. Few publications were from developing nations. There was limited stakeholder involvement in the VSP designs and a lack of robust research designs, consistent outcome measures, conceptual underpinnings, and immersive technologies. Despite this, promising trends for student experience, knowledge, communication, and critical thinking skills using VSPs have emerged.

**Conclusion:** This review maps the VSP evidence across medicine, nursing, midwifery and allied health. Before a systematic review is feasible across healthcare, allied health and midwifery research require greater representation. Based on the highlighted gaps, other areas for future research are suggested.

**WHAT IS ALREADY KNOWN ON THIS TOPIC:**

- Digital placements in undergraduate nursing and medicine have been studied in one existing systematic review, providing evidence that learning outcomes for knowledge and practice were equivalent to traditional placements.
- VSPs are a subset of digital placements that are computer-generated. With the increasing trend towards VSPs, an updated scoping review across a wider range of professions was justified.

**WHAT THIS STUDY ADDS:**

- Scoping the literature on VSPs across healthcare for undergraduate and postgraduate students, provides a map across professions, specialities, countries, designs, content, and outcomes.

**HOW THIS STUDY MIGHT AFFECT RESEARCH, PRACTICE OR POLICY:**

- Gaps in allied health and midwifery VSP research highlight populations of focus. Future VSPs should consider Interprofessional Education (IPE) and resource sharing with developing countries. The benefits of immersive technologies are yet to be considered, and improvements to VSP design and research methodology are recommended.

## Introduction

Practice placements are important activities in the training of healthcare students. They promote the application of knowledge to a practical setting for developing the skills, attitudes and behaviours expected of a healthcare professional [1–3]. Placements allow active involvement in care delivery under supervision, and the opportunity to receive feedback on student performance [4]. In other words, student learning on placement is contextualised to future practice.

Simulation-based education is an alternative to traditional practice placements. In traditional placements, students enter a workplace and learn through observation and participation in actual clinical events. In contrast, healthcare simulation is a technique that produces a scenario designed to represent a real-life practice situation for experiential learning [5–6]. Compared with traditional placements, simulation can ensure that low-frequency and high-risk cases or situations receive sufficient practice in a safe space, without mistakes causing harm to real persons [7]. Thus, the advantage of simulation is the ability to control and direct case-based learning.

With technological advances, simulation-based education has expanded into virtual environments. Technology innovation accelerated during the pandemic, and healthcare training must keep pace with continued advances [8–9]. The increasing complexity of healthcare, with developments in science and technology, will require the future workforce to be agile, lifelong learners, with the ability to substitute skills across professions [10–12]. Using virtual simulated environments could provide students with the opportunity to support these aims.

A VSP is defined in this research as a computer-generated version of a practice placement. Considering the importance of practice placements, the advantages of simulation-based learning and the recent advances in technology, this topic is relevant for review.

### Background for the Scoping Review

A preliminary search of MEDLINE, the Cochrane Database of Systematic Reviews and Joanna Briggs Institute Evidence Synthesis was conducted on 17^th^ June 2022 to locate any existing or underway reviews on the topic. A systematic review [13] was identified, focused on digital placements for undergraduate nursing and medical students. The review also included non-computer-generated experiences such as telemedicine and on-screen role-play rather than being restricted to VSPs. Whilst sixteen studies were located in their search in April 2021, the increased trend towards implementing computer-generated placements (VSPs) within undergraduate and postgraduate programmes across the wider health professions justifies the current review.

As VSPs are an emerging field, mapping the literature across healthcare and analysing gaps are recommended before more specific research questions are defined [14–16]. Therefore, a scoping review method was selected to conduct a broader search across medical, nursing, midwifery, and allied healthcare, for undergraduate and postgraduate students who undertook VSPs. The objective is to assess the range and types of evidence related to VSPs, across the healthcare professions. It was hypothesised that mapping the literature regarding VSPs across healthcare might highlight innovations in one speciality that could be applied to another. Sufficient research in a specific area may underline the requirement for a systematic review, and conversely, gaps in the literature could justify new research areas.

## Methods

This study followed the stages detailed in a framework for scoping reviews [14]:

1. Identify the research question
2. Identify relevant studies
3. Study selection
4. Charting the data
5. Collating, summarising, and reporting the results

An a-priori protocol used the Joanna Briggs Institute template for scoping reviews [16] and was registered with the Open Science Framework (DOI 10.17605/OSF.IO/AY5GH) [17]. The PRISMA-ScR checklist (see online supplemental file A1) ensured methodological rigour when reporting this review [18].

### Review Questions

1. What is the scope of the literature relating to VSPs for healthcare students?
2. What outcomes are reported in relation to the students undertaking VSPs?

### Relevant Studies

The eligibility criteria are tabled below using the SPIDER [19] and PCC [16] formats:

The selection criteria were piloted by screening 50 titles and abstracts. This process generated 94% agreement between two reviewers (JS & MG) and served to clarify the selection criteria. In discussion with a 3rd reviewer (NT), the Health and Care Professions Council (HCPC) definition for allied health [20] was adopted in place of the NHS criteria [21]. This was decided because the HCPC definition includes practitioner psychologists. It was reasoned that psychology students might form a population well suited to VSPs, with the treatment emphasis on talking therapies.

### Search strategy

An initial limited search of MEDLINE and CINAHL was undertaken on 28^th^ June 2022 to identify articles on the topic. The text words contained in the titles and abstracts of relevant articles and index terms were used to develop a full search strategy. This was checked by a healthcare research librarian and run on MEDLINE on 3^rd^ August 2022 (see online supplemental file A2). The search strategy was then adapted for each database. The databases searched included MEDLINE, CINAHL, AMED, Cochrane Database, PsychINFO, ERIC, SCOPUS, ScienceDirect and Biomed Central. Grey literature sources include PubMed, EThOS, ProQuest (dissertations), Google Scholar and IEEE Xplore. Searches were limited to English language and dated from 1^st^ January 2020. The date limitation was justified given that VSP research has essentially emerged post-pandemic.

Supplementary search strategies were employed using existing knowledge and networks, contacting relevant organisations, hand searching journals and checking the reference list of all included sources and relevant reviews. Advances in Simulation, British Medical Journal: Simulation and Technology Enhanced Learning (BMJ STEL) and Clinical Simulation in Nursing were hand searched. These supplementary searches were conducted by one reviewer (JS) and checked by another (NT).

An updated database search was conducted on the 9^th^ of July 2023. A second reviewer (MG) checked the title/abstract and full text selection decisions. Registries (Clinical_Trials.gov WHO ICTRP and the Cochrane Database) were searched for additional papers [25]. Updated hand searches were performed in Advances in Simulation and Clinical Simulation in Nursing (BMJ STEL had since discontinued). A second reviewer (NT) checked these supplementary searches.

### Source Selection

Following the database searches, all identified citations were uploaded into Endnote [22], and duplicates were removed. Each potential duplicate was confirmed separately, rather than using batch automation to prevent the removal of false positives [23]. The citations were exported to Rayyan and re-checked for any missed duplicates [24].

Once the pilot screening was complete, the remaining titles/abstracts were screened independently by two reviewers against the revised criteria, and potentially relevant sources were retrieved in full text. These were assessed in detail against the inclusion criteria by two independent reviewers (JS & MG), blinded in Rayyan. 100% agreement was reached between the reviewers through discussion. Further details of the source selection, including a list of references excluded at full text screening are detailed in online supplemental file A3.

### Data Charting

A Microsoft Excel spreadsheet was used as a data charting tool to standardise obtaining information from the papers. Two independent reviewers conducted a pilot of five included papers to assess the utility of the information charted and generate emerging themes. Consensus was reached between reviewers (JS & MG) on the charting method, and modifications were made to the spreadsheet, to improve the quality of charted data (see online supplemental file A4). Following this, one reviewer (JS) charted the remaining data, which was checked by another (NT).

A table of included study characteristics was collated, and numerical analysis in Microsoft Excel was undertaken to provide descriptive statistics. The size of the data set was manageable enough to organise findings across the PAGER domains (patterns, advances, gaps, evidence for practice and research recommendations) [26], for synthesis, without the use of NVIVO software (as was planned in the protocol).

## Results

The search results and selection process are reported in the Preferred Reporting Items for Systematic Reviews and Meta-analyses (PRISMA) flow diagram (Figure 1):

**Figure 1:**
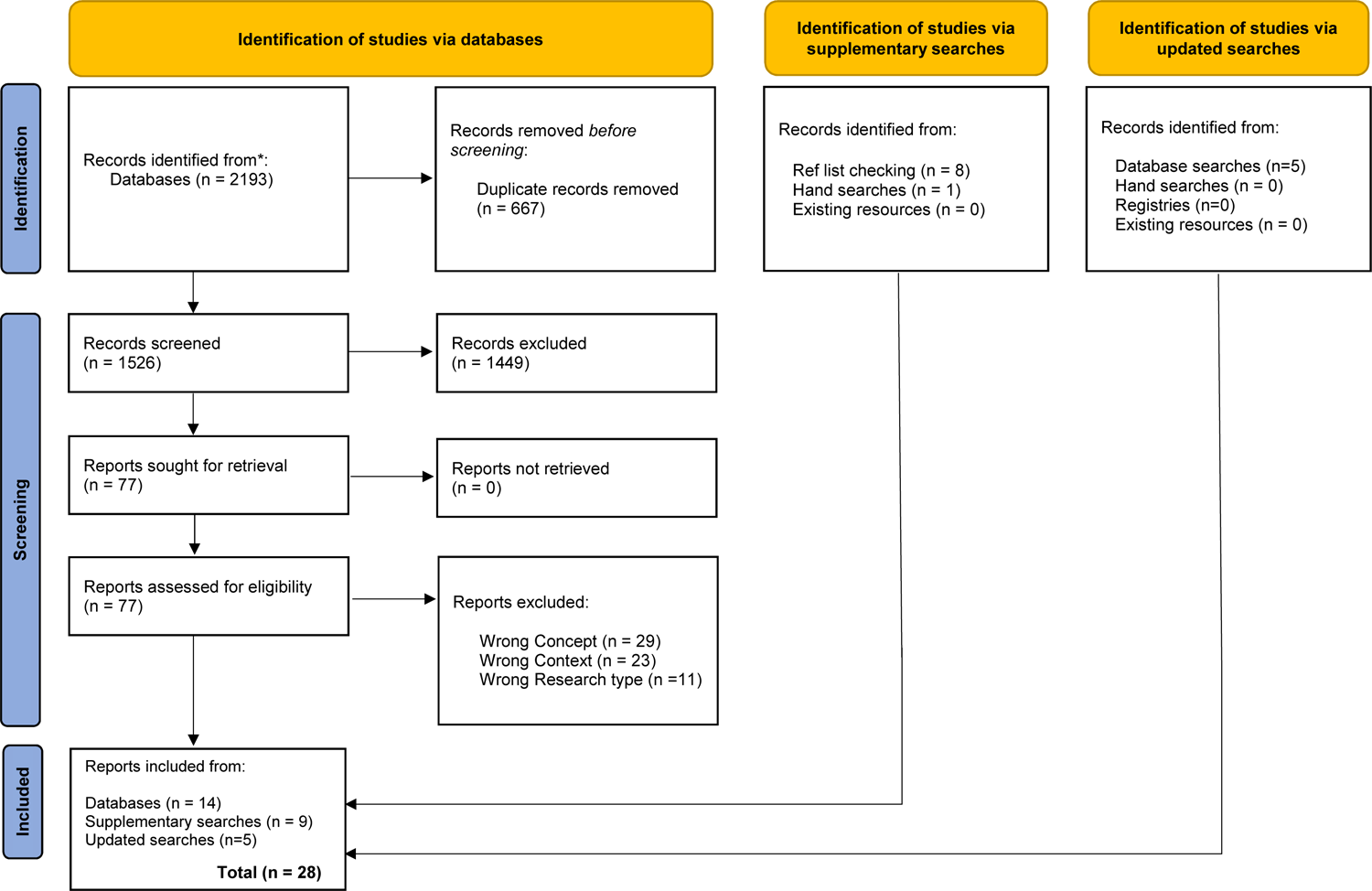
PRISMA chart. Modified from [27]

The characteristics of the twenty-eight included papers are summarised in online supplemental file A5 and the PAGER themes are summarised in Table 2:

**Table 1:**
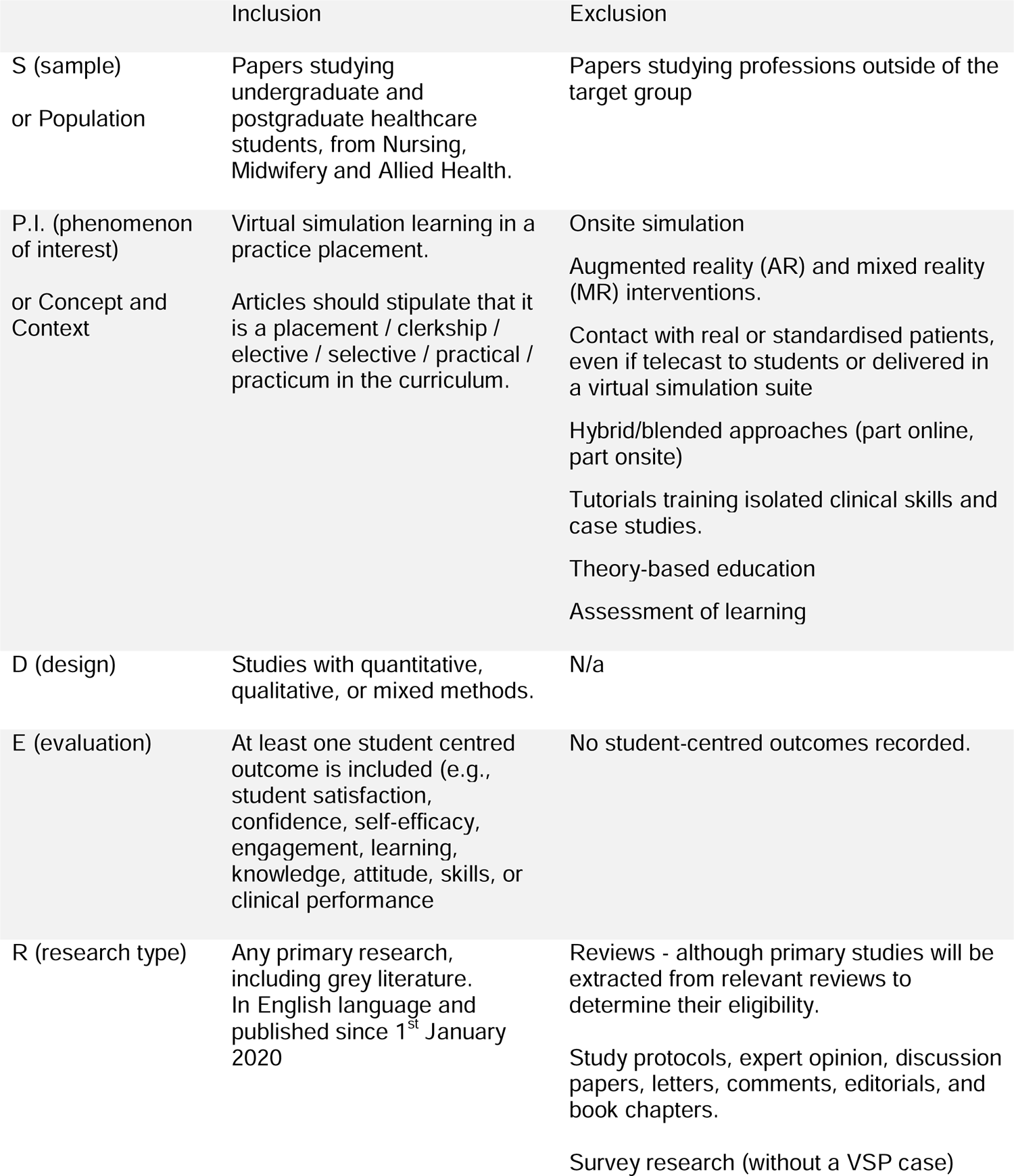
Eligibility criteria in PCC and SPIDER formats.

|  | Inclusion | Exclusion |
| --- | --- | --- |
| S (sample)<br>or Population | Papers studying undergraduate and postgraduate healthcare students, from Nursing, Midwifery and Allied Health. | Papers studying professions outside of the target group |
| P.I. (phenomenon of interest)<br>or Concept and Context | Virtual simulation learning in a practice placement.<br>Articles should stipulate that it is a placement / clerkship / elective / selective / practical / practicum in the curriculum. | Onsite simulation<br>Augmented reality (AR) and mixed reality (MR) interventions.<br>Contact with real or standardised patients, even if telecast to students or delivered in a virtual simulation suite<br>Hybrid/blended approaches (part online, part onsite)<br>Tutorials training isolated clinical skills and case studies.<br>Theory-based education<br>Assessment of learning |
| D (design) | Studies with quantitative, qualitative, or mixed methods. | N/a |
| E (evaluation) | At least one student centred outcome is included (e.g., student satisfaction, confidence, self-efficacy, engagement, learning, knowledge, attitude, skills, or clinical performance) | No student-centred outcomes recorded. |
| R (research type) | Any primary research, including grey literature. In English language and published since 1<sup>st</sup> January 2020 | Reviews - although primary studies will be extracted from relevant reviews to determine their eligibility.<br>Study protocols, expert opinion, discussion papers, letters, comments, editorials, and book chapters.<br>Survey research (without a VSP case) |

**Table 2:** PAGER Framework Themes Summary.

| Patterns | Advances | Gaps | Evidence for Practice | Research Recommendations |
| --- | --- | --- | --- | --- |
| Publications from developed countries (see Figure 2) | Innovations occurred mostly in countries with resources to support VSP development | Few publications from the developing world | VSPs can be delivered remotely and are scalable (useful for supporting training in the developing world) | Sharing resources across countries and overcoming barriers such as internet connectivity or access to devices |
| Narrow profession focus | All VSPs occurred within single profession silos | No Interprofessional education (IPE) | Support for VSPs delivering on improved discipline specific skills | The development of IPE VSPs to train collaborative capabilities |
|  | Student populations were mostly medical / nursing | No allied health representation |  | Allied health groups could inform future IPE VSPs |
| Pandemic Response | Rapid innovation to shift from in-person placement to VSPs in response to COVID-19 restrictions. | Research planned under time pressure may explain the lack of robust experimental design and conceptual frameworks | Positive outcomes suggest that VSPs could be utilised beyond the pandemic response | With less time pressure, future research could consider conceptual frameworks, with more robust experimental designs |
| Stakeholder involvement in the VSP design | Most studies involved university faculty. Others also included clinicians. | Few incorporated student input and consultation. No evidence of co-creation. | Design that involves student participation throughout the process better serves the end user needs | Participatory research designs should include all stakeholders, including students and service users (who ultimately benefit) |
| Use of generic platforms and screen-based delivery | Platforms such as Microsoft Teams, Zoom and existing Learning Management Systems (LMS) were used to facilitate VSP delivery | Limited use of bespoke healthcare education software or use of virtual reality (VR). No headsets or haptics. No conversational Artificial Intelligence (AI). | Student feedback frequently rated the live interaction with facilitators positively | Future research could test live interaction with bespoke VR software. Headsets and haptic research may emerge as devices become more ubiquitous |
| A focus on case-based learning | VSPs were oriented towards clinical cases and the development of knowledge, clinical reasoning, decision making and communication | Practical skills training was rare. Few featured social determinants of health / community interventions | Evidence for improved knowledge, clinical thinking, and communication skills from VSP interventions | Hybrid approaches are currently more suitable for practical skills but haptics in VR may feature as technology improves. Community interventions link well with IPE |
| Survey based outcome measures | Most VSPs were evaluated through custom designed surveys and student marks | Few validated outcome measure scales or standardised examinations | Evaluations were overall positive and improvements in test scores were equivalent to in-person cohorts | Validated outcome measures and standardised tests in future trials would provide more robust data for meta-analysis |

Key patterns and gaps are mapped across all included studies in Table 3:

**Table 3:** Key patterns and gaps.

| Citation | Patterns |  |  |  | Gaps |  |  |  |  |  |  |  |
| --- | --- | --- | --- | --- | --- | --- | --- | --- | --- | --- | --- | --- |
|  | Developed Nation | Medical or Nursing Profession | Pandemic Response | Generic Software | Population |  | Experimental Design |  | Students involved in the design | Conceptual Frameworks | Software<br>Bespoke software | Hardware<br>VR equipment |
|  |  |  |  |  | IPE | Allied Health | Comparator group | Pre & post measures |  |  |  |  |
| [28] | ✓ | ✓ | ✓ | ✓ |  |  | ✓ |  |  |  |  |  |
| [29] | ✓ | ✓ | ✓ | ✓ |  |  |  |  |  | ✓ |  |  |
| [30] | ✓ | ✓ | ✓ | ✓ |  |  |  |  |  | ✓ | ✓ |  |
| [31] | ✓ | ✓ | ✓ | ✓ |  |  |  |  |  |  | ✓ |  |
| [32] | ✓ | ✓ | ✓ | ✓ |  |  |  |  |  |  | ✓ |  |
| [33] | ✓ | ✓ | ✓ | ✓ |  |  | ✓ |  |  | ✓ |  |  |
| [34] |  |  | ✓ | ✓ |  |  |  | ✓ |  | ✓ |  |  |
| [35] | ✓ | ✓ | ✓ | ✓ |  |  |  |  |  |  | ✓ |  |
| [36] |  | ✓ | ✓ | ✓ |  |  |  |  |  |  |  |  |
| [37] | ✓ | ✓ | ✓ | ✓ |  |  |  | ✓ | ✓ |  |  |  |
| [38] | ✓ | ✓ | ✓ | ✓ |  |  |  |  |  |  | ✓ |  |
| [39] | ✓ | ✓ | ✓ | ✓ |  |  |  | ✓ |  | ✓ |  |  |
| [40] | ✓ | ✓ | ✓ | ✓ |  |  |  |  |  | ✓ | ✓ |  |
| [41] |  | ✓ | ✓ | ✓ |  |  |  | ✓ |  | ✓ | ✓ |  |

Table 3: Key patterns and gaps
| Citation<br>(cont'd) | Patterns |  |  |  | Gaps |  |  |  |  |  |  |  |
| --- | --- | --- | --- | --- | --- | --- | --- | --- | --- | --- | --- | --- |
|  | Developed<br>Nation | Medical<br>or Nursing<br>Profession | Pandemic<br>Response | Generic<br>Software | Population |  | Experimental Design |  | Students<br>involved in<br>the design | Conceptual<br>Frameworks | Software<br>Bespoke<br>software | Hardware<br>VR<br>equipment |
|  |  |  |  |  | IPE | Allied<br>Health | Comparator<br>group | Pre & post<br>measures |  |  |  |  |
| [42] | ✓ | ✓ | ✓ | ✓ |  |  |  |  |  |  |  |  |
| [43] | ✓ | ✓ | ✓ | ✓ |  |  |  |  |  | ✓ |  |  |
| [44] | ✓ | ✓ | ✓ | ✓ |  |  |  |  | ✓ |  | ✓ |  |
| [45] | ✓ | ✓ | ✓ | ✓ |  |  | ✓ |  |  | ✓ | ✓ |  |
| [46] | ✓ | ✓ | ✓ | ✓ |  |  |  |  |  | ✓ | ✓ |  |
| [47] | ✓ | ✓ | ✓ | ✓ |  |  |  |  |  |  | ✓ |  |
| [48] | ✓ | ✓ | ✓ | ✓ |  |  |  | ✓ | ✓ |  |  |  |
| [49] | ✓ |  | ✓ | ✓ |  | ✓ |  |  |  | ✓ | ✓ |  |
| [50] | ✓ | ✓ | ✓ | ✓ |  |  |  | ✓ | ✓ | ✓ | ✓ |  |
| [51] | ✓ | ✓ | ✓ | ✓ |  |  | ✓ |  |  |  | ✓ |  |
| [52] | ✓ | ✓ | ✓ | ✓ |  |  |  |  |  | ✓ | ✓ |  |
| [53] | ✓ | ✓ | ✓ | ✓ |  |  |  |  |  |  | ✓ |  |
| [54] | ✓ | ✓ | ✓ | ✓ |  |  |  | ✓ |  | ✓ |  |  |
| [55] |  | ✓ | ✓ | ✓ |  |  | ✓ |  |  | ✓ | ✓ |  |

Most of the included papers were published in developed countries. The global distribution of publications is illustrated (in Figure 2).

**Figure 2:**
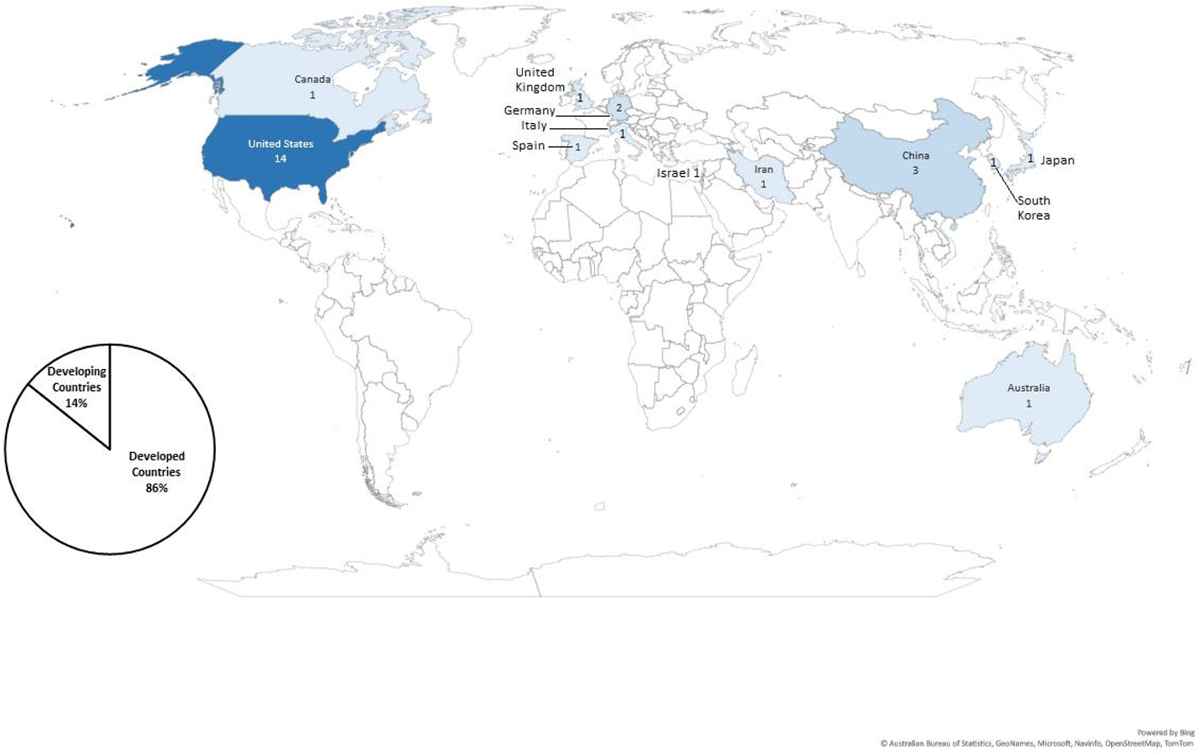
Country of origin of included papers

### Range of Professions

The literature was predominantly medical and nursing research, constituting 93% of the included papers. The distribution by profession and breakdowns by specialty are illustrated in online supplemental file A6. Diagnostic radiology rotations were the most prevalent VSPs in Medicine, and Paediatrics in Nursing.

### Pandemic Response

All the VSPs in the included papers were developed in response to COVID-19 restrictions, which discontinued face to face (FTF) practice placements.

### Experimental Designs

The most basic study design was a single group, with a post-intervention measure, featuring in sixteen papers. Seven papers compared measures pre- and post-intervention. Five papers compared VSP outcomes to a previous cohort of students who completed FTF placements pre-pandemic.

### Stakeholder involvement

Practice partners (clinicians working in practice) were involved in the course development with faculty in eight studies [29, 32, 37, 41, 44, 48, 50, 55] and students were involved in four. Three studies developed a needs assessment from student surveys [34, 43, 52]. None involved service users.

### Conceptual Frameworks

Conceptual underpinnings include pedagogy, theoretical frameworks, and professional standards. Although no single paper covered all elements, underpinning concepts are evident across the literature, summarised in online supplemental file A7.

### Software

All studies used generic software such as Zoom or Microsoft Teams for screen-based communication, and many used existing learning management systems to host files and activities. Others adopted commercial software applications, allowing students to conduct a history by selecting from a menu of interview questions. None used conversational AI (computer-generated conversation, assisted by artificial intelligence). Some applications presented VR patient avatars with which the student could direct a physical examination, although this was delivered via a screen [31, 38, 40–41, 51] and one study provided an interactive community setting in screen-based VR [53]. All software resources are outlined in online supplemental file A8.

### Intended learning outcomes

The focus of most VSPs were clinical cases, through which knowledge, clinical reasoning, decision making, and communication skills were developed. Skill learning was generally visualised through virtual patient encounters and instructional/walk-through procedure videos. The social determinants of health were the focus in two studies [50, 53] and another facilitated student in teaching roles [42].

### Outcomes

The most common outcome measures were custom developed student evaluation questionnaires, followed by exam marks. Custom questionnaires provided positive feedback for student experience, satisfaction, and usability, although some technical issues and zoom fatigue were cited [31, 50]. Three papers reported a 100% pass rate on their VSPs [35, 49, 52], and four used a standardised exam to demonstrate comparable outcomes with FTF cohorts [45, 51], or the national average [30, 32].

Table 4 summarises the outcomes of research that employed a repeated measures design or group comparisons.

**Table 4:**
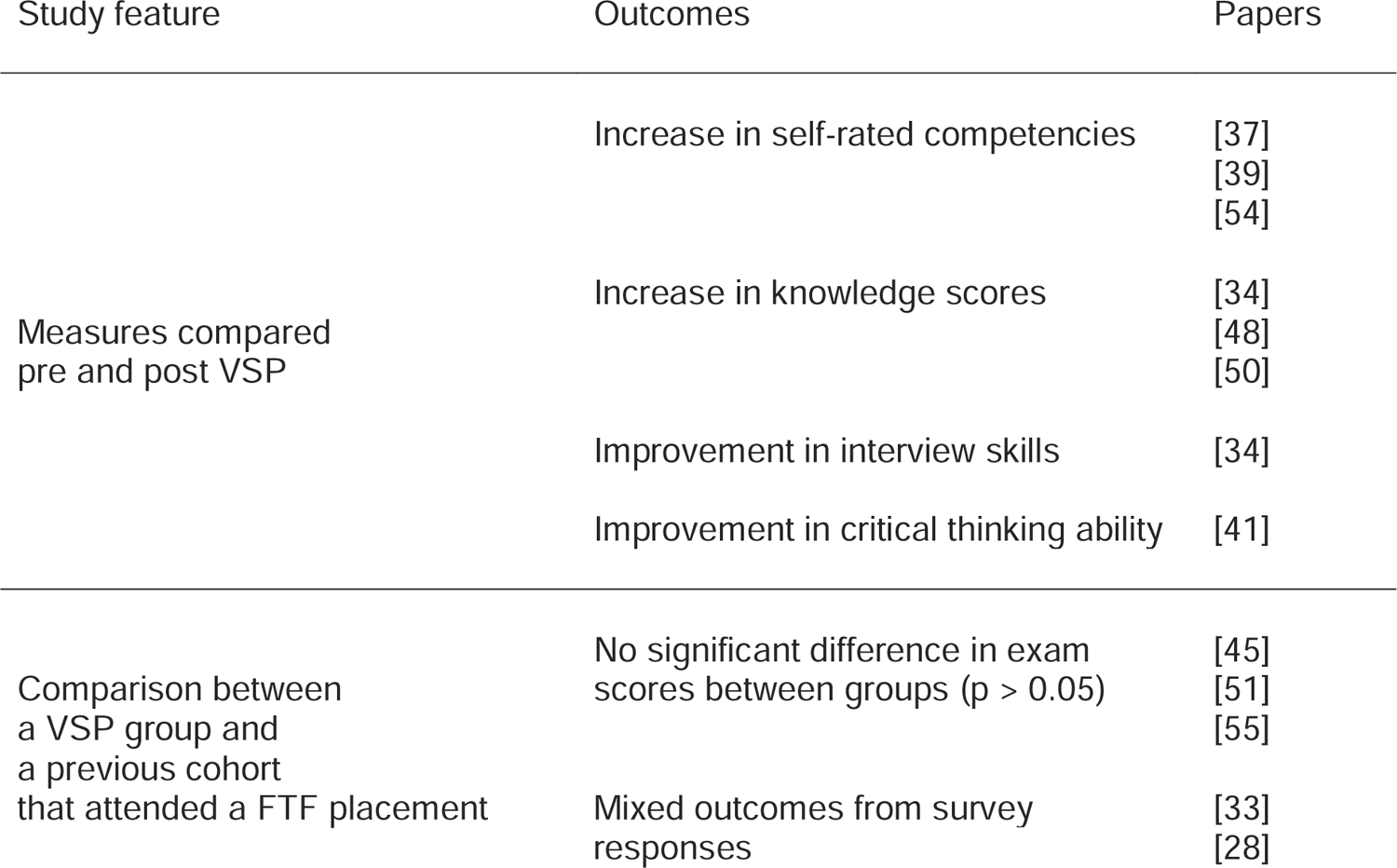
Outcomes from intra/inter group comparisons.

| Study feature | Outcomes | Papers |
| --- | --- | --- |
| Measures compared pre and post VSP | Increase in self-rated competencies | [37]<br>[39]<br>[54] |
|  | Increase in knowledge scores | [34]<br>[48]<br>[50] |
|  | Improvement in interview skills | [34] |
|  | Improvement in critical thinking ability | [41] |
| Comparison between a VSP group and a previous cohort that attended a FTF placement | No significant difference in exam scores between groups ($p > 0.05$) | [45]<br>[51]<br>[55] |
|  | Mixed outcomes from survey responses | [33]<br>[28] |

When measures were compared pre- and post-VSP, there was a trend of improvement in self-rated competencies, knowledge scores and critical thinking skills. However, when the comparison is made with traditional FTF placements, the pattern is less clear. There were no differences in grades when post-VSP exam scores were compared with previous cohorts’ who attended a FTF placement pre-pandemic. Student satisfaction was comparable in a study conducted in medical general practice, but professional exchange and learning scored higher in the VSP, while the attainment of new skills and attitudes scored higher in the FTF placement [33]. One paper compared students who participated in virtual readouts (the radiology equivalent of patient rounds) with students who attended workplace readouts pre-pandemic [28]. The educational value was comparable in survey results, though students on the VSP rated slightly higher for perceived interaction. That FTF students were mostly observing on their placement might explain this finding. Conversely, FTF students had greater confidence in using the workstations, considered the case because the VSP students were unable to operate PACS workstations remotely.

## Discussion

### Outcomes and Research designs

The pattern of positive student evaluation, improvement from baseline measures post VSP, and equivalence in exam scores, compared with in-person cohorts, appears promising. However, these results should be interpreted with caution. The objective of a scoping review is to map the literature for patterns and gaps, rather than in-depth appraisal of the quality of the papers. The findings compare with a systematic review that examined digital clinical education more broadly [13]. Standalone digital education was reported to be as effective as conventional learning for knowledge and practice, in nursing and medicine. However, there are some methodological concerns with this systematic review [13]. There was no a-priori protocol, and the study lacked a pilot to test the methods. A librarian’s involvement in verifying the search strategy was not reported, grey literature was not searched, and duplicate processes were absent for the study selection and data extraction stages.

There are several barriers to conducting a systematic review of VSPs across healthcare. Firstly, there is insufficient research across midwifery and allied health [34,49]. Another consideration is that all student evaluations in this scoping review were custom designed by the authors. Therefore, the inconsistency of outcome measures might prevent meaningful comparison across the papers. One study used previously researched scales for clinical thinking ability, academic self-efficacy, and student engagement, which demonstrated good reliability [41]. Some of the exams were standardised [30, 32, 45, 51], but none compared the baseline marks of each group to determine whether there were differences at the outset. In all cases, VSP exam scores were compared with a previous cohort that attended placement FTF pre-pandemic, or the national average, rather than adopting a prospective design. It is clear from the paucity of research outside nursing and medicine, the lack of prospective research designs and inconsistent, non-validated outcome measures, that research into VSPs is in its infancy.

### VSP design and Stakeholder involvement

Elements of thoughtful VSP design are evident across several papers. Frameworks such as ADDIE (analysis, design, development, implementation, and evaluation), ensure that there is structure to the process, and stakeholder needs are met [30]. Existing curricula [54–55], or processes such as Kern’s 6-step model for curricular development could be used [43, 45, 46, 50, 52]. If framed within existing standards [40, 49], VSPs can align with specified learning outcomes. Principles in pedagogy such as andragogy [29, 30] and online learning [33, 50], ensure that VSPs build features that engage students with experiential learning [30], promote problem solving [29–30, 39] and active reflection [49]. The conceptual underpinnings documented across this body of literature could provide a blueprint for best practice in VSP design.

Stakeholder involvement is a key process to inform the design of a VSP. Service users could inform the content and students are the end users of a VSP, yet no service user involvement was documented, and students were involved in a minority of studies. When they were involved, student surveys informed a needs assessment, or they were consulted early in the process. This is a tokenistic approach compared with co-creation, the preferred method of engaging with stakeholders. Co-creation involves a collective effort with all stakeholders to collaborate across the entire design, development, implementation, and testing phases [57]. A UK university provided an overview of VSP development within a nursing programme, which included students, service users and other universities throughout [58]. Their working group comprised of academics, clinicians, a service user and carer involvement lead and an education technology lead. Therefore, in addition to underpinning VSP design with the relevant conceptual frameworks (pedagogical principles, theoretical frameworks, and published standards), broad stakeholder co-creation is optimal.

### Interprofessional Education (IPE)

VSPs have the potential to break down silos between professions, by delivering IPE over a virtual platform. IPE is defined as two or more professions, ‘learning with from and about one another to improve collaborative practice and quality of care’ [59], p4. The intended outcome is to improve mutual understanding, teamwork, and leadership among different professionals [60]. VSPs have advantages over FTF training in building asynchronous activities for flexibility in timetabling and hosting synchronous activities without geographical constraints [61]. Given the relevance of IPE to quality care and the fit with virtual technologies, IPE-VSPs may be an important area for future research.

### Content and Technologies

Disciplines that rely on image-based diagnoses are more easily adapted to screen-based delivery, and consistent with this, diagnostic radiology, and pathology VSPs together constitute over 20% of the medical papers in this review. In the development of this scoping review, the researchers anticipated that psychology might be suited to VSPs due to the nature of talking-based therapies over physical skills, although it is possible that psychological presentations were considered too complex to portray accurately in computer-generated simulations. With future developments in conversational AI and the growing acceptance of this technology, this situation may change. Similarly, professions that rely heavily on hands-on assessment, such as physiotherapy, may feature more in virtual reality spaces when improvements in haptic technology emerge. In the meantime, virtual placements that require complex conversation are likely to include telecast or telemedicine simulations. Likewise, virtual placements that teach advanced handling skills might adopt a hybrid or blended approach. Selecting studies that conducted a computer-generated placement entirely online, rather than employing a hybrid or blended approach may explain why all papers in this review were pandemic responses. COVID-19 necessitated a rapid shift to provide VSPs as a replacement for lost clinical hours [62]. However, these VSPs were often produced in a short timeframe, under emergency situations, and may explain why few papers featured robust experimental designs and conceptual frameworks. Further, hybrid or blended approaches could combine the strengths of both virtual simulation and FTF approaches.

Replacing FTF placement hours with simulation is a contentious issue. Accordingly, a Delphi study considered the benefits and limitations of this approach [63]. Expert consensus across multiple professions agreed that between 11 and 30% of hours replaced would be acceptable, and this aligns with the current allocation set by the Nursing and Midwifery Medical Council [64]. VSPs in the curriculum may offset some pressure on workplace settings as they attempt to fulfil the NHS long-term plan [11]. However, this does not negate the need to continue building workplace placement capacity [63]. VSPs can be considered an additional pedagogy that offers a different, yet complimentary experience to traditional FTF placements.

### Strengths and Weaknesses

The strengths of this study relate to the methodology. A structured process for defining search terms was undertaken, and a librarian was consulted for the search strategy. A range of databases were searched across medical and technology specialities. Grey literature sources were searched, and an updated search included trial registries. An a-priori protocol was registered, and a subset of data was piloted to determine the declared changes. Duplicate processes in study selection and data charting were employed, and existing guidelines were used to design the protocol, synthesise the findings and report the paper.

The weaknesses relate to limiting the search to English language. This increases the risk of language bias, and a search unrestricted by language may have yielded a more balanced global representation. Whilst the limited number of publications from developing countries could be a function of the language limitation, it is also likely that countries with greater resources were better positioned to make the rapid shift to online education and publish their research during a global health emergency. Virtual platforms are suited to sharing resources and overcoming geographical constraints to access expertise, and VSPs present an opportunity to address inequality in healthcare education moving forward.

## Conclusion

The emerging trends for VSPs in this review demonstrate some positive outcomes, although a systematic review would be required to quality assess and evaluate the evidence across healthcare education. For this to be possible, VSP research from allied health and midwifery, require greater representation. Specific outcome measures for this new mode of learning also need to be developed and tested, thus ensuring that valid, reliable, and consistent measures are used across future studies. Future research should include prospective designs with repeated measures and control/comparator groups to strengthen the evidence. This review highlights the need for VSP design to be co-created with a wider range of stakeholders and underpinned by pedagogical principles, theoretical frameworks, and published standards. Research into student engagement using VR headsets, haptics, and conversational AI in VSPs, is another area for future research as technologies develop. The pandemic has revealed an opportunity to augment placement capacity through VSPs. There is the potential for future VSPs to feature IPE, thus promoting joined-up care in healthcare graduates. There is also the opportunity for VSPs to improve local and global access to quality clinical education experiences.

## Supporting information

Supplemental File A3 Mono

Supplemental File A3 Colour

Supplemental File A6 Mono

## Data Availability

All data produced in the present work are contained in the manuscript

## Acknowledgements

We acknowledge our health librarian, Carlo Avillo, for assisting with the search strategy. This review will contribute towards a PhD award for J.S.

## Conflicts of interest

None

### Appendices (in the Supplemental Material)

Appendix 1: PRISMA-ScR Checklist

Appendix 2: MEDLINE Search strategy

Appendix 3: Screening Decisions

Appendix 4: Revised data charting tool

Appendix 5: Table of included study characteristics

Appendix 6: Papers by Professions

Appendix 7: Conceptual Frameworks

Appendix 8: Bespoke healthcare technology

