## Supplemental File A1 for "Virtual Simulated Placements in Healthcare Education: A scoping review"

Supplementary File A1: PRISMA-ScR Checklist [18]

| Section | Item | PRISMA-ScR Checklist Item | Reported on page # |
|---|---|---|---|
| **Title** | 1 | Identify the report as a scoping review | 1 |
| **Abstract** | | | |
| Structured summary | 2 | Provide a structured summary that includes (as applicable) background, objectives, eligibility criteria, sources of evidence, charting methods, results, and conclusions that relate to the review questions and objectives. | 1 |
| **Introduction** | | | |
| Rationale | 3 | Describe the rationale for the review in the context of what is already known. Explain why the review questions or objectives lend themselves to a scoping review approach. | 2-3 |
| Objectives | 4 | Provide an explicit statement of the questions and objectives being addressed with reference to their key elements (for example, population or participants, concepts, and context) or other relevant key elements used to conceptualize the review questions or objectives. | 3 |
| **Methods** | | | |
| Protocol and registration | 5 | Indicate whether a review protocol exists; state if and where it can be accessed (for example, a Web address); and if available, provide registration information, including the registration number. | 3 & 18 |
| Eligibility criteria | 6 | Specify characteristics of the sources of evidence used as eligibility criteria (for example, years considered, language, and publication status), and provide a rationale. | 4-5 |
| Information sources | 7 | Describe all information sources in the search (for example, databases with dates of coverage and contact with authors to identify additional sources), as well as the date the most recent search was executed. | 5 |
| Search | 8 | Present the full electronic search strategy for at least 1 database, including any limits used, such that it could be repeated. | Suppl. file A2 |
| Selection of Sources of Evidence | 9 | State the process for selecting sources of evidence (that is, screening and eligibility) included in the scoping review. | 5 |
| Data charting process | 10 | Describe the methods of charting data from the included sources of evidence (for example, calibrated forms or forms that have been tested by the team before their use, and whether data charting was done independently or in duplicate) and any processes for obtaining and confirming data from investigators. | 6 |
| Data items | 11 | List and define all variables for which data were sought and any assumptions and simplifications made. | Suppl. file A4 |
| Critical Appraisal of Individual Sources of Evidence | 12 | *Optional*: If done, provide a rationale for conducting a critical appraisal of included sources of evidence; describe the methods used and how this information was used in any data synthesis (if appropriate). | Not done. Reasons on p14 |
| Summary Measures | 13 | *Not Relevant:* This item from the original PRISMA is not applicable for scoping reviews because a meta-analysis is not done (that is, summary measures are not relevant). | Not done |

| | | | |
|---|---|---|---|
| Synthesis of Results | 14 | Describe the methods of handling and summarizing the data that were charted. | 6 |
| Risk of bias across studies | 15 | *Not Applicable:* This item from the original PRISMA is not applicable for scoping reviews because the scoping review method is not intended to be used to critically appraise (or appraise the risk of bias of) a cumulative body of evidence. | Not done |
| Additional Analyses | 16 | *Not Applicable:* This item from the original PRISMA is not applicable for scoping reviews because additional analyses, including sensitivity or subgroup analyses and meta-regression, are not done. | Not done |
| **Results** | | | |
| Selection of Sources of Evidence | 17 | Give numbers of sources of evidence screened, assessed for eligibility, and included in the review, with reasons for exclusions at each stage, ideally using a flow diagram. | 7 |
| Characteristics of Sources of Evidence | 18 | For each source of evidence, present characteristics for which data were charted and provide the citations. | 9-10 |
| Critical Appraisal Within Sources of Evidence | 19 | *Optional:* If done, present data on critical appraisal of included sources of evidence (see item 12). | Not done |
| Results of Individual Sources of Evidence | 20 | For each included source of evidence, present the relevant data that were charted that relate to the review questions and objectives. | Suppl. file A5 |
| Synthesis of Results | 21 | Summarise or present the charting results as they relate to the review questions and objectives. | 8 |
| Risk of Bias Across Studies | 22 | *Not applicable:* This item is not applicable for scoping reviews. See explanation for item 15. | Not done |
| Additional Analyses | 23 | *Not applicable:* This item is not applicable for scoping reviews. See explanation for item 16 | Not done |
| **Discussion** | | | |
| Summary of Evidence | 24 | Summarise the main results (including an overview of concepts, themes, and types of evidence available), link to the review questions and objectives, and consider the relevance to key groups. | 14-15 |
| Limitations | 25 | Discuss the limitations of the scoping review process. | 15-16 |
| Conclusions | 26 | Provide a general interpretation of the results with respect to the review questions and objectives, as well as potential implications or next steps. | 16 |
| **Funding** | 27 | Describe sources of funding for the included sources of evidence, as well as sources of funding for the scoping review. Describe the role of the funders of the scoping review. | 16 |
