## Supplemental File A2 for "Virtual Simulated Placements in Healthcare Education: A scoping review"

Supplementary File A2: Search Strategy (MEDLINE)
(Run on 3rd August 2022)

| Search ID # | Query | Results |
|---|---|---|
| S5 | Limited to date from January 2020, English language and Humans | 880 |
| S4 | S1 AND S2 AND S3 | 3,013 |
| S3 | (TI Clinical education OR AB Clinical education OR TI Clinical Placement* OR AB Clinical Placement* OR TI Placement* OR AB Placement* OR TI Practical* OR AB Practical* OR TI Practicum OR AB Practicum OR TI Clerkship* OR AB Clerkship*) OR (MH "Interdisciplinary Placement") | 549,959 |
| S2 | ( TI Virtual Simulat* OR AB Virtual Simulat* OR TI Virtual reality OR AB Virtual reality OR TI Virtual patient OR AB Virtual patient OR TI Virtual Environment OR AB Virtual Environment OR TI Online OR AB Online OR TI Remote OR AB Remote ) OR ( (MH "Virtual Reality”) OR (MH "Computer Simulation") ) | 498,694 |
| S1 | ( TI Healthcare student* OR AB Healthcare student* OR TI Health care student* OR AB Health care student* OR TI Medic* student* OR AB Medic* student* OR TI Nurs* student* OR AB Nurs* student* OR TI Midwi* student* OR AB Midwi* student* OR TI Allied health student* OR AB Allied health student* ) OR (MH “Students, Health Occupations”) | 132,641 |
