## Supplemental File A3 Mono for "Virtual Simulated Placements in Healthcare Education: A scoping review"

### Supplementary File A3: Screening Decisions

#### Initial Search:

##### Database Records Breakdown

| Databases | Results |
|---|---|
| AMED | 1 |
| BioMed Central | 2 |
| CINAHL | 159 |
| Cochrane | 125 |
| ERIC | 6 |
| EthOS | 2 |
| Google Scholar | 80 |
| IEEE Xplore | 264 |
| MEDLINE | 880 |
| ProQuest Dissertations | 15 |
| PsychINFO | 32 |
| PubMed | 611 |
| Science Direct | 11 |
| Scopus | 5 |
| Total | 2193 |

Database search papers screened out at full text stage, with reasons for exclusion

| Reference | Reason | Comments |
|---|---|---|
| Addis, B., et al. (2022). "Virtual elective placements for medical students during COVID-19." Medical Education **56**(5): 576-577. | Wrong Research Type | Commentary piece. Also consisted of observing cases over video conferencing. No outcome measures reported. |
| Aster, A., et al. (2022). "Use of a Serious Game to Teach Infectious Disease Management in Medical School: effectiveness and Transfer to a Clinical Examination." Frontiers in medicine **9**. | Wrong Concept | The aim was to compare it to an OSCE and see if it could substitute - so it's about assessment |
| Betts, L., et al. (2020). "Using Virtual Interactive Digital Simulator to Enhance Simulation Experiences for Undergraduate Nursing Students." Nursing education perspectives 41(3): 193-194. | Wrong Concept | Patient simulator in a touch-table format for simulations in a centre |
| Bradford, H. M., et al. (2021). "Rapid Curricular Innovations During COVID-19 Clinical Suspension: Maintaining Student Engagement with Simulation Experiences." Journal of midwifery & women's health 66(3): 366-371. | Wrong Research Type | There were several innovations but not one set VSP described - aside from the OSCEs, many of the activities were optional and could substitute for some clinical hours. |
| Cárdenas-Cruz, A., et al. (2022). "An example of adaptation: experience of virtual clinical skills circuits of internal medicine students at the Faculty of Medicine, University of Granada (Spain) during the COVID-19 pandemic." Med Educ Online **27**(1): 2040191. | Wrong Concept | Was a combination of classroom work and clinical skill circuits (equivalent to OSCE style set up) |
| Castro, M. R. H., et al. (2021). "Lessons From Learners: Adapting Medical Student Education During and Post COVID-19." Academic medicine : journal of the Association of American Medical Colleges 96(12): 1671-1679. | Wrong Concept | The placements weren't simulations, but real patient contact facilitated over telemedicine |
| Chircop, A. and S. Cobbett (2020). "Gett'n on the bus: evaluation of Sentinel CityÂ®3.0 virtual simulation in community/population health clinical placement." International Journal of Nursing Education Scholarship **17**(1). | Wrong Concept | The agency/neighbourhood component of the placement was replaced with VSim, but all subjects continued the concurrent parts of their training |
| Ctri (2021). "Clinical virtual simulation is effective in nursing education." https://trialsearch.who.int/Trial2.aspx?TrialID=CTRI/2021/01/030826. | Wrong Research Type | This is a protocol for a future study |

| Reference | Reason | Comments |
| --- | --- | --- |
| DePietro, D. M., et al. (2021). "Medical Student Education During the COVID-19 Pandemic: Initial Experiences Implementing a Virtual Interventional Radiology Elective Course." Academic Radiology 28(1): 128-135. | Wrong Concept | Discussions were based on real - rather than simulated patients |
| Elsayes, K. M., et al. (2021). "Multidisciplinary Approach in Teaching Diagnostic Radiology to Medical Students: The Development, Implementation, and Evaluation of a Virtual Educational Model." Journal of the American College of Radiology : JACR 18(8): 1179-1187. | Wrong Context | Wasn't a placement - it was a series of 10 case based, 1 hr lectures |
| Feeley, A., et al. (2021). "Bedside teaching from a distance; surgical education in the COVID era." British journal of surgery **108**: vi231-. | Wrong Concept | Not a simulation - remote interaction with the hospital patients was facilitated through smart glasses |
| Feeley, A., et al. (2022). "Student Acceptance of Virtual Bedside Surgical Tutorials During COVID-19; A Randomized Control Trial." Journal of Surgical Research 270: 261-265. | Wrong Concept | Not a simulation - remote interaction with the hospital patients was facilitated through smart glasses - appears to be the same study as the conference proceedings by the same author |
| Foohey, S., et al. (2022). "Developing the Virtual Resus Room: Fidelity, Usability, Acceptability, and Applicability of a Virtual Simulation for Teaching and Learning." Academic medicine : journal of the Association of American Medical Colleges **97**(5): 679-683. | Wrong Context | The focus was on the VRR and there was mention of the 2 simulation cases being integrated into a VSP but no detail on it |
| Goldenson, R. P., et al. (2022). "The Virtual Homeroom: Utility and Benefits of Small Group Online Learning in the COVID-19 Era." Current problems in diagnostic radiology 51(2): 152-154. | Wrong Concept | This is the same study as Durfee (included) - but the focus was on the homerooms within the clerkship |
| Grady, Z. J., et al. (2022). "From the Operating Room to Online: Medical Student Surgery Education in the Time of COVID-19." The Journal of surgical research 270: 145-150. | Wrong Context | A 2 week non-clinical elective to prepare and generate interest in surgical placements |
| Guerandel, A., et al. (2021). "An approach to teaching psychiatry to medical students in the time of Covid-19." Ir J Psychol Med **38**(4): 293-299. | Wrong Concept | The online module replaces 2 of the 6 weeks of the placement |
| Gummerson, C. E., et al. (2021). "Broadening learning communities during COVID-19: developing a curricular framework for telemedicine education in neurology." BMC medical education **21**(1): 549. | Wrong Concept | The virtual rounds were based on real rather than simulated patients, with the opportunity to read their notes and/or have a telemedicine interview before the lessons |

| Reference | Reason | Comments |
|---|---|---|
| He, Z. and C. Xiong (2021). Design and Application of Rehabilitation Psychology Practical Teaching System Based on VR Technology. | Wrong Context | Replaced a traditional mode without practical operation due to lack of resources to rehab disabled patients - so this was a university module rather than a placement |
| Hwang, G.-J., et al. (2022). "The effectiveness of the virtual patient-based social learning approach in undergraduate nursing education: A quasi-experimental study." Nurse education today 108: 105164. | Wrong Context | Examined the use of Virtual patients in a social learning framework and compared it to traditional leaching for 1 week - not a placement |
| Killam, L. A. and M. Luctkar-Flude (2021). "Virtual Simulations to Replace Clinical Hours in a Family Assessment Course: Development Using H5P, Gamification, and Student Co-Creation." Clinical Simulation in Nursing 57: 59-65. | Wrong Concept | In person component |
| Kullberg, M. L., et al. (2020). "E-learning to improve suicide prevention practice skills among undergraduate psychology students: Randomized controlled trial." JMIR Mental Health 7(1). | Wrong Context | e-learning module on suicidal behaviour - Not a placement |
| Lakhtakia R. Virtual Microscopy in Undergraduate Pathology Education: An early transformative experience in clinical reasoning. Sultan Qaboos Univ Med J. 2021 Aug;21(3):428-435. doi: 10.18295/squmj.4.2021.009. Epub 2021 Aug 29. PMID: 34522409; PMCID: PMC8407892. | Wrong Concept | conducted as a hybrid placement (computer lab) |
| Larraga-GarcÃ¬a, B., et al. (2021). "Design and Development of an Interactive Web-Based Simulator for Trauma Training: A Pilot Study." Journal of Medical Systems 45(11): 96. | Wrong Context | Single case virtual simulation |
| Margolin, E. J., et al. (2021). "Reimagining the Away Rotation: A 4-Week Virtual Subinternship in Urology." Journal of surgical education 78(5): 1563-1573. | Wrong Concept | includes a teleconferenced clinic with real patients, so not a simulation |
| Middeke, A., et al. (2020). "Transfer of Clinical Reasoning Trained With a Serious Game to Comparable Clinical Problems: a Prospective Randomized Study." Simulation in healthcare 15(2): 75-81. | Wrong Context | The emphasis was on whether clinical reasoning skills learned through the game could be applied in subsequent patient cases - not a placement |
| Mikhail, D., et al. (2021). "Changing the Status Quo: Developing a Virtual Sub-Internship in the Era of COVID-19." Journal of surgical education 78(5): 1544-1555. | Wrong Context | Not a placement, but a guidebook |
| Miller, A. and K. Guest (2021). "Rising to the Challenge: The Delivery of Simulation and Clinical Skills during COVID-19." Comprehensive child and adolescent nursing 44(1): 6-14. | Wrong Research Type | Discussion paper |

| Reference | Reason | Comments |
|---|---|---|
| Pal, A. D., et al. (2022). "Virtual Simulation for Advanced Practice Registered Nurse Students: Adapting to Shortage of Clinicals." Journal for Nurse Practitioners 18(5): 563-568. | Wrong Context | Virtual simulations credited 8 FTF placement hours |
| Ramjan, F., et al. (2021). "Psychiatry through a Screen: Adapting Training for a New Reality?" Psychiatria Danubina 33: 18-21. | Wrong Research Type | Commentary summarising curricular changes and student experiences but no specific VSP |
| Ray, J., et al. (2021). “Virtual Telesimulation for Medical Students During the COVID-19 Pandemic” Academic medicine : journal of the Association of American Medical Colleges 96(10) 1431-1435 | Wrong Concept | Telecast activities from an in-person simulation - so not computer generated |
| Roberts, M. L. and J. O. E. Mazurak (2022). "Virtual Clinical Experiences in Nursing Education: Applying a Technology-Enhanced Storyboard Technique to Facilitate Contextual Learning in Remote Environments." Nursing education perspectives 43(4): 260-261. | Wrong Research Type | Storyboard technique for design detailed rather than a specific placement |
| Rosenthal, H. B., et al. (2021). "A Near-Peer Educational Model for Online, Interactive Learning in Emergency Medicine." Western Journal of Emergency Medicine: Integrating Emergency Care with Population Health 22(1): 130-135. | Wrong Context | Lecture series |
| Roskvist, R., et al. (2020). "Provision of e-learning programmes to replace undergraduate medical students' clinical general practice attachments during COVID-19 stand-down." Education for primary care : an official publication of the Association of Course Organisers, National Association of GP Tutors, World Organisation of Family Doctors 31(4): 247-254. | Wrong Research Type | General overview of their model - not empirical |
| Rydel, T. A., et al. (2021). "Hands Off Yet All In: A Virtual Clerkship Pilot in the Ambulatory Setting During the COVID-19 Pandemic." Academic medicine : journal of the Association of American Medical Colleges 96(12): 1702-1705. | Wrong Concept | Live rather than simulated telehealth sessions in the placement |
| Safdieh, J. E., et al. (2021). "Curricular response to COVID-19: real-time interactive telehealth experience (RITE) program." Medical education online 26(1): 1918609. | Wrong Concept | Live telehealth rather than simulated patients |
| Salem, J., et al. (2020). "Virtual Patient Journey: a novel learning resource." The clinical teacher 17(3): 315-319. | Wrong Context | 3 x 8 min videos of an asthma patient's journey - not a placement |

| Reference | Reason | Comments |
| --- | --- | --- |
| Santo, L. D., et al. (2022). "The emotional side of the e-learning among nursing students: The role of the affective correlates on e-learning satisfaction." Nurse Educ Today 110: 105268. | Wrong Research Type | Survey examining emotions and the cognitive and social dimensions of e-learning |
| Seymour-Walsh, A. E., et al. (2020). "Practical approaches to pedagogically rich online tutorials in health professions education." Rural Remote Health 20(2): 6045. | Wrong Research Type | Guidelines regarding designing online tutorials with sound pedagogical underpinnings |
| Shoemaker, M. M., et al. (2021). "Novel application of telemedicine and an alternate EHR environment for virtual clinical education: A new model for primary care education during the SARS-CoV-2 pandemic." International Journal of Medical Informatics 153: 104526. | Wrong Concept | Live telehealth rather than simulated patients |
| Smith, N., et al. (2021). "A mixed methods evaluation of the Paediatric Musculoskeletal Matters (PMM) online portfolio." Pediatr Rheumatol Online J 19(1): 85. | Wrong Context | e-learning analytics and a survey evaluation of online modules / app for paeds MSK Ax - not a placement |
| Spear, L. G., et al. (2022). "Rethinking Clinical Trial Radiology Workflows and Student Training: Integrated Virtual Student Shadowing Experience, Education, and Evaluation." Journal of digital imaging 35(3): 723-731. | Wrong Context | Online resource to improve image measurement and reporting - not a placement |
| Srinivasan, S. L. and O. Burton (2021). "Is It Possible to Replicate a Surgical Placement for Medical Undergraduates Virtually in Response to the COVID-19 Pandemic?" Journal of the American College of Surgeons **233**(5): e170-. | Wrong Research Type | Only an abstract - Contacted the author (via Researchgate) to retrieve the full study but no reply |
| Streitlein-Böhme, I., et al. (2021). "We can also do online - evaluation of the accompanying digital seminar of the elective subject "General Practice" during intership (PJ) at Ruhr-University Bochum." GMS J Med Educ **38**(4): Doc73. | Wrong Concept | The online seminars were part of the placement - a FTF clinical had taken place prior |
| Svoboda, S. A., et al. (2021). "Inspired by COVID-19 isolation: Evolving educational techniques in dermatology residency programs." Clinics in dermatology **39**(1): 41-44. | Wrong Research Type | Commentary regards telemedicine in dermatology clinical education |
| Teichgräber, U., et al. (2021). "Virtual inverted classroom to replace in-person radiology lectures at the time of the COVID-19 pandemic - a prospective evaluation and historic comparison." BMC medical education 21(1): 611. | Wrong Context | Physical converted to a virtual flipped classroom - not a placement |

| Reference | Reason | Comments |
|---|---|---|
| Tinoco, J. D. S., et al. (2021). "Effect of educational intervention on clinical reasoning skills in nursing: A quasi-experimental study." Nurse education today 105: 105027-. | Wrong Concept | Assessed an app for developing clinical reasoning skills - not a placement and also mentions a face-face stage |
| Twogood, R., et al. (2020). "Rapid implementation and improvement of a virtual student placement model in response to the COVID-19 pandemic." BMJ open quality **9**(4). | Wrong Concept | Live telehealth rather than simulated patients |
| UrbanovÃ¡, E., et al. (2022). "Virtual patients: an option for future distance midwifery education?" International Journal of Nursing Education Scholarship 19(1). | Wrong Context | Detailed the design of a set of virtual patients but it was not conducted as a placement |
| Vielsmeier, V., et al. (2020). "Digital teaching with interactive case presentations of ENT diseases - discussion of utilisation and motivation of students." GMS J Med Educ **37**(7): Doc100. | Wrong Concept | Has the option of an in-person component |
| Wade, S. W. T., et al. (2020). "Adaptive tutorials versus web-based resources in radiology: a mixed methods analysis in junior doctors of efficacy and engagement." BMC medical education **20**(1): 303. | Wrong Context | Compared adaptive tutorials to web based learning in graduate doctors - not a placement |
| Walsh, H., et al. (2022). "Innovative Hospital-Based Pediatric Virtual Learning for Nursing Students." Nurse Educator 47(2): E30-E33. | Wrong Context | Not a placement - but some participants were able to credit 1-2 hours in place of FTF |
| Wands, L., et al. (2020). "Positive Outcomes of Rapid Freeware Implementation to Replace Baccalaureate Student Clinical Experiences." The Journal of nursing education 59(12): 701-704. | Wrong Concept | One resource used was augmented reality but the rest were virtual |
| Wang, M., et al. (2022). "Intelligent virtual case learning system based on real medical records and natural language processing." BMC Med Inform Decis Mak **22**(1): 60. | Wrong Context | Documents the process of combining AI natural language processing and electronic notes - intended for an adjunct to clinical learning rather than a stand-alone placement |
| Waugh, S., Devin, J., Lam, A.KY. et al. E-learning and the virtual transformation of histopathology teaching during COVID-19: its impact on student learning experience and outcome. BMC Med Educ 22, 22 (2022). https://doi.org/10.1186/s12909-021-03066-z | Wrong Context | The intervention is replacing a classroom lesson (lectures & seminars). No mention of placement |
| Weber, A. M., et al. (2021). "An outpatient telehealth elective for displaced clinical learners during the COVID-19 pandemic." BMC medical education **21**(1): 174. | Wrong Concept | Live telehealth rather than simulated patients |

| Reference | Reason | Comments |
| --- | --- | --- |
| Weiss, M. E., et al. (2021). "Effectiveness of using a simulation combined with online learning approach to develop discharge teaching skills." Nurse education in practice **52**: 103024. | Wrong Concept | Intervention was focused on improving discharge planning - not a placement |
| Wendt, S., et al. (2021). "A virtual COVID-19 ophthalmology rotation." Survey of ophthalmology **66**(2): 354-361. | Wrong Concept | Included in person or video conferenced real patients |
| Wiese, L. K., et al. (2021). "Responding to a simulated disaster in the virtual or live classroom: Is there a difference in BSN student learning?" Nurse education in practice **55**: 103170. | Wrong Context | Single emergency disaster simulation - comparing live and virtual versions - not a placement |
| Williams, J., et al. (2022). "Development of a simulation placement in a pre-registration nursing programme." British journal of nursing (Mark Allen Publishing) **31**(10): 549-554. | Wrong Context | Blended |
| Winship, J. M., et al. (2020). "A case study in rapid adaptation of interprofessional education and remote visits during COVID-19." Journal of Interprofessional Care **34**(5): 702-705. | Wrong Concept | Live patient contact via telecoms rather than simulated patients |
| Wittenberg, E., et al. (2021). "COVID 19-transformed nursing education and communication competency: Testing COMFORT educational resources." Nurse education today **107**: 105105. | Wrong Context | Educational model to support COVID-19 related communication skills - not a placement |
| Wu, Y., et al. (2021). "Flipping the Passive Radiology Elective by Including Active Learning." Canadian Association of Radiologists journal = Journal l'Association canadienne des radiologistes **72**(4): 621-627. | Wrong Concept | Not virtual |
| Yan, H., et al. (2021). Research on Real-time Medical Online Learning Content Recommendation based on Multi-view Data Mining. | Wrong Concept | Data mining and analysis across MOOC platforms - not relevant |

#### Updated Search

#### Database Records Breakdown

| Databases | Results |
|---|---|
| AMED | 0 |
| BioMed Central | 1 |
| CINAHL | 63 |
| Cochrane | 77 |
| ERIC | 3 |
| EthOS | 1 |
| Google Scholar | 88 |
| IEEE Xplore | 199 |
| MEDLINE | 394 |
| ProQuest Dissertations | 3 |
| PsychINFO | 7 |
| PubMed | 279 |
| Science Direct | 8 |
| Scopus | 0 |
| Total | 1123 |

PRISMA Chart for Updated Search

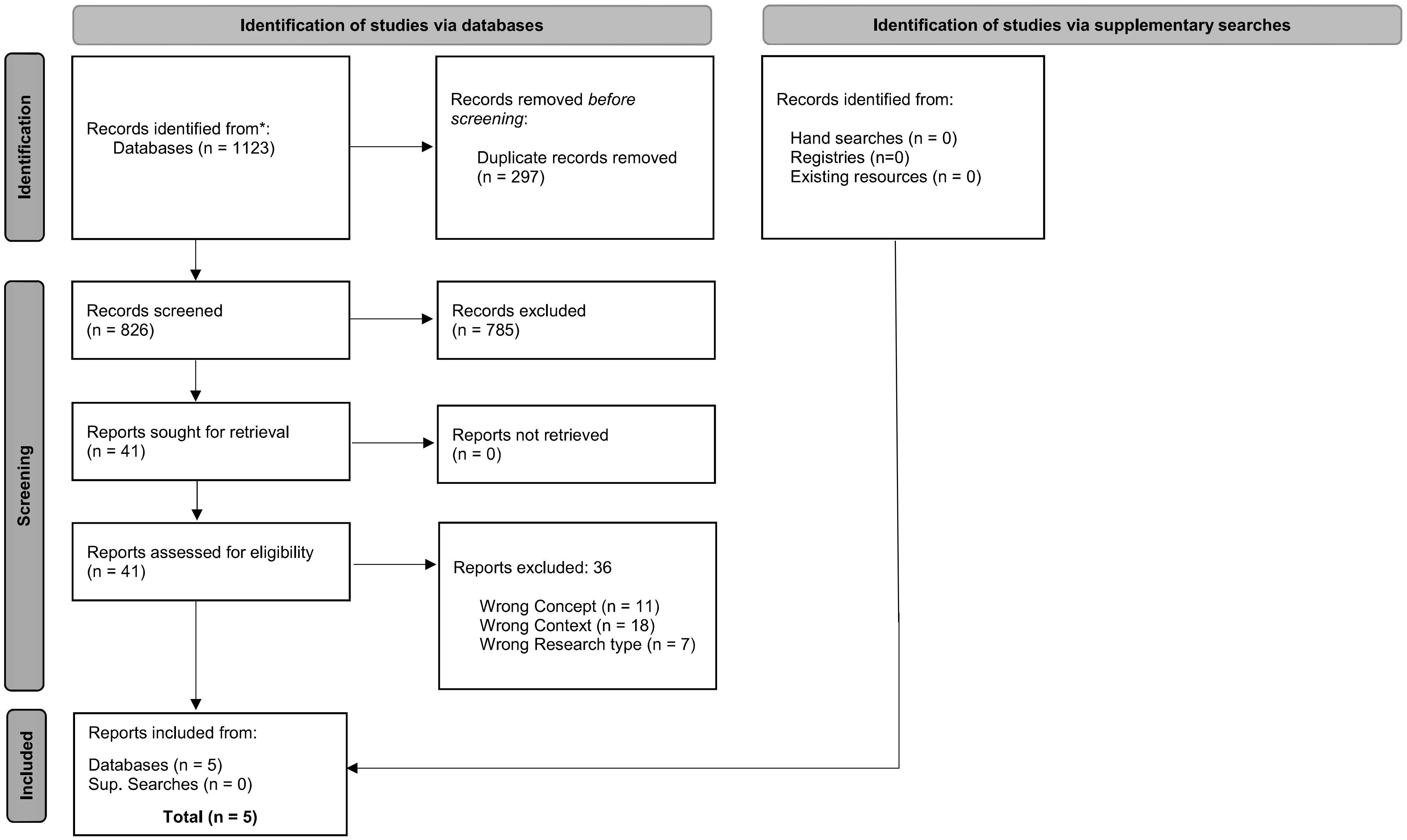


*Modified From:* Page MJ, McKenzie JE, Bossuyt PM, Boutron I, Hoffmann TC, Mulrow CD, et al. The PRISMA 2020 statement: an updated guideline for reporting systematic reviews. BMJ 2021;372:n71. doi: 10.1136/bmj.n71. For more information, visit: http://www.prisma-statement.org/

Database search papers screened out at full text stage, with reasons for exclusion

| **Reference** | **Reason** | **Comment** |
|---|---|---|
| Aubrey, C., et al. (2022). "Using simulated placements to promote inclusive practices: a higher education institution faculty's experience of delivery." | wrong study type | Selected abstracts |
| Chan, P. P., et al. (2023). "Flipped Classroom Case Learning vs Traditional Lecture-Based Learning in Medical School Ophthalmology Education: a Randomized Trial." Academic medicine. | wrong concept | There is a component of FTF / classroom work |
| Cieslowski, B., et al. (2023). "The Development and Pilot Testing of Immersive Virtual Reality Simulation Training for Prelicensure Nursing Students: A Quasi-Experimental Study." Clinical Simulation in Nursing 77: 6-12. | wrong concept | IVR group also participated in clinical cases, so hybrid |
| Duffy B, Tully R, Stanton AV. An online case-based teaching and assessment program on clinical history-taking skills and reasoning using simulated patients in response to the COVID-19 pandemic. BMC Med Educ. 2023 Jan 4;23(1):4. doi: 10.1186/s12909-022-03950-2. PMID: 36600232; PMCID: PMC9811710. | wrong concept | Use of telecast simulated patients |
| Elnaga, H. H. A., et al. (2023). "Virtual versus paper-based PBL in a pulmonology course for medical undergraduates." BMC medical education 23(1): 433. | wrong context | 2 clinical cases in one module - not a placement |
| Hardin, L. M., et al. (2022). "Innovative Team Approach for Achieving DNP Program Competencies for Distance Learners During the COVID-19 Pandemic." Nursing education perspectives 43(5): 318-320. | wrong study type | Report without any measurement of student outcomes |
| Henze, S. M., et al. (2022). "Digital adaptation of teaching disaster and deployment medicine under COVID-19 conditions: a comparative evaluation over 5 years." BMC medical education 22(1): 717. | wrong context | A course rather than a placement |
| Herrmann-Werner, A., et al. (2022). "Where there are challenges, there are opportunities: An undergraduate medical students' teaching concept for mental health in times of COVID-19." PloS one 17(11): e0277525. | wrong context | Teaching course converted online: Not a placement |

| Reference | Reason | Comment |
| --- | --- | --- |
| Ijaz, A., et al. (2023). "Online learning: an effective option for teaching ENT to medical students?" The Journal of laryngology and otology 137(5): 560-564. | wrong context | Not a placement |
| Khanittanuphong, P., et al. (2022). "The impact of the transition from flipped classroom to online lectures on learning outcomes and student satisfaction in a rehabilitation medicine clerkship during the COVID-19 pandemic." BMC medical education 22(1): 885. | wrong context | Comparing the flipped classroom component of a clerkship and it's switch to online lectures - not a placement |
| Kumar, P. R., et al. (2022). "F1-taught orthopaedic teaching programme for students (FOTS)." Postgraduate Medical Journal 98(1163): 710-717. | wrong context | 6 x 60 min weekly joint assessment session - not a placement |
| Liaw, S. Y., et al. (2023). "Effectiveness of an online program using telesimulation for academic–clinical collaboration in preparing nurse preceptors' roles." Journal of Clinical Nursing (John Wiley & Sons, Inc.) 32(7): 1115-1124. | wrong concept | Students role played the simulated telemedicine session in VR over zoom |
| Liu, C., et al. (2023). "Beyond Skin Deep: case-based online modules to teach multidisciplinary care in dermatology among clerkship students." BMC medical education 23(1): 90. | wrong context | An online pre-clerkship for a FTF clerkship - not a placement |
| Lubinski, B. and A. Weinstein (2022). "Working together but physically siloed-telehealth and interprofessional education among medicine clerkship students." Journal of general internal medicine 37: S662. | wrong study type | Selected abstract |
| McAllister, L. L., et al. (2022). "A descriptive case report of telesupervision and online case-based learning for speech and language therapy students in Vietnam during the COVID-19 pandemic." The South African journal of communication disorders = Die Suid-Afrikaanse tydskrif vir Kommunikasieafwykings 69(2): e1-e6. | wrong concept | Live patient sessions were broadcast - not computer generated |
| Mergen, M., et al. (2023). "Immersive training of clinical decision making with AI driven virtual patients - a new VR platform called medical tr.AI.ning." GMS journal for medical education 40(2): Doc18. | wrong study type | Research plan |
| Meyer, E. G., et al. (2023). "The Effectiveness of Online Experiential Learning in a Psychiatry Clerkship." Acad Psychiatry 47(2): 181-186. | wrong concept | Use of simulated patients over telecoms - not computer generated |

| Reference | Reason | Comment |
| --- | --- | --- |
| O'Connor, M. and L. Rainford (2023). "The impact of 3D virtual reality radiography practice on student performance in clinical practice." Radiography (London, England : 1995) 29(1): 159-164. | wrong concept | 7 hours VR practice within a 4 week placement |
| Omlor, A. J., et al. (2022). "Comparison of immersive and non-immersive virtual reality videos as substitute for in-hospital teaching during coronavirus lockdown: a survey with graduate medical students in Germany." Med Educ Online 27(1): 2101417. | wrong context | Observation replaced - but not a placement |
| Petersen, K., et al. (2023). "Online Virtual Patient Cases vs. Weekly Classroom Lectures in an Internal Medicine Clerkship: Effects on Military Learner Outcomes." Military Medicine 188(5): 914-920. | wrong context | The Virtual Patient was a substitute for lectures. Didactic content - Not a Placement |
| Salje, J. and M. Moyo (2023). "Implementation of a virtual student placement to improve the application of theory to practice." British journal of nursing (Mark Allen Publishing) 32(9): 434-441. | wrong concept | One day teaching on campus (hybrid) |
| Schoenherr, D. T., et al. (2022). "Development and evaluation of an online integrative histology module: simple design, low-cost, and improves pathology self-efficacy." Medical education online 27(1): 2011692. | wrong context | Course rather than a placement |
| Shen, A. H., et al. (2022). "Designing a Plastic and Reconstructive Surgery Virtual Curriculum: Assessment of Medical Student Knowledge, Surgical Skill, and Community Building." Plastic and reconstructive surgery 150(3): 691-700. | wrong study type | Selected abstract |
| Shin, H. and D. Rim (2023). "Development and assessment of a curriculum model for virtual simulation in nursing: curriculum development and pilot-evaluation." BMC medical education 23(1): 284. | wrong context | Not a placement (yet) |
| Shorey, S., et al. (2023). "Evaluation of a Theory-Based Virtual Counseling Application in Nursing Education." Computers, informatics, nursing : CIN 41(6): 385-393. | wrong context | Not a placement |
| Showstark, M., et al. (2023). "Results and lessons learned from a virtual multi-institutional problem-based interprofessional learning approach: The VIPE program." Journal of Interprofessional Care 37(1): 164-167 | wrong context | Not a placement |

| Reference | Reason | Comment |
|---|---|---|
| Su, J., et al. (2023). "Effects of a virtual simulation-based interprofessional education activity for rehabilitation nursing using shared resources: A quasi-experimental study." Nurse education today 126: 105832. | wrong context | Not a placement - but an activity |
| Thirsk, L. M., et al. (2022). "Effect of online versus in-person clinical experiences on nursing student's competency development: A cross-sectional, quasi-experimental design." Nurse education today 117: 105461. | wrong study type | Focus on the surveys rather than description of the online placement |
| Uraiby, H., et al. (2022). "Fostering intrinsic motivation in remote undergraduate histopathology education." Journal of clinical pathology 75(12): 837-843. | wrong concept | The facilitator acted as a virtual patient on screen - not computer generated / virtual simulation |
| Uzun Aksoy, M., et al. (2023). "The effectiveness of the using scenario and video in distance nursing education during COVID-19 pandemic." Teaching & Learning in Nursing 18(1): 24-29. | wrong context | Not a placement |
| Varachotisate, P., et al. (2023). "Student academic performance in non-lecture physiology topics following the abrupt change from traditional on-site teaching to online teaching during COVID-19 pandemic." Medical education online 28(1): 2149292. | wrong context | Pre-clerkship theory focussed teaching - not a placement |
| Vives, M., et al. (2023). "Teaching Psychotherapy to Psychiatric/Mental Health Nurse Practitioner Students in the Virtual Classroom." Issues in Mental Health Nursing 44(6): 551-561. | wrong context | Skills based course: Not a placement |
| Vogt, L., et al. (2022). "Telemedicine in medical education: An example of a digital preparatory course for the clinical traineeship - a pre-post comparison." GMS journal for medical education 39(4): Doc46. | wrong concept | Simulated (standardised) patients are used over telemedicine - not computer generated |
| Wicks, S. K., et al. (2023). "Anaesthetic National Teaching Programme for Students (ANTPS)." Postgraduate Medical Journal 99(1172): 613-623. | wrong context | A course, not a placement |
| Wojniusz, S., et al. (2022). "Active digital pedagogies as a substitute for clinical placement during the COVID-19 pandemic: the case of physiotherapy education." BMC medical education 22(1): 843. | wrong concept | A FTF history and assessment were included as an activity |
