## Supplemental File A3 Colour for "Virtual Simulated Placements in Healthcare Education: A scoping review"

PRISMA Chart for Updated Search

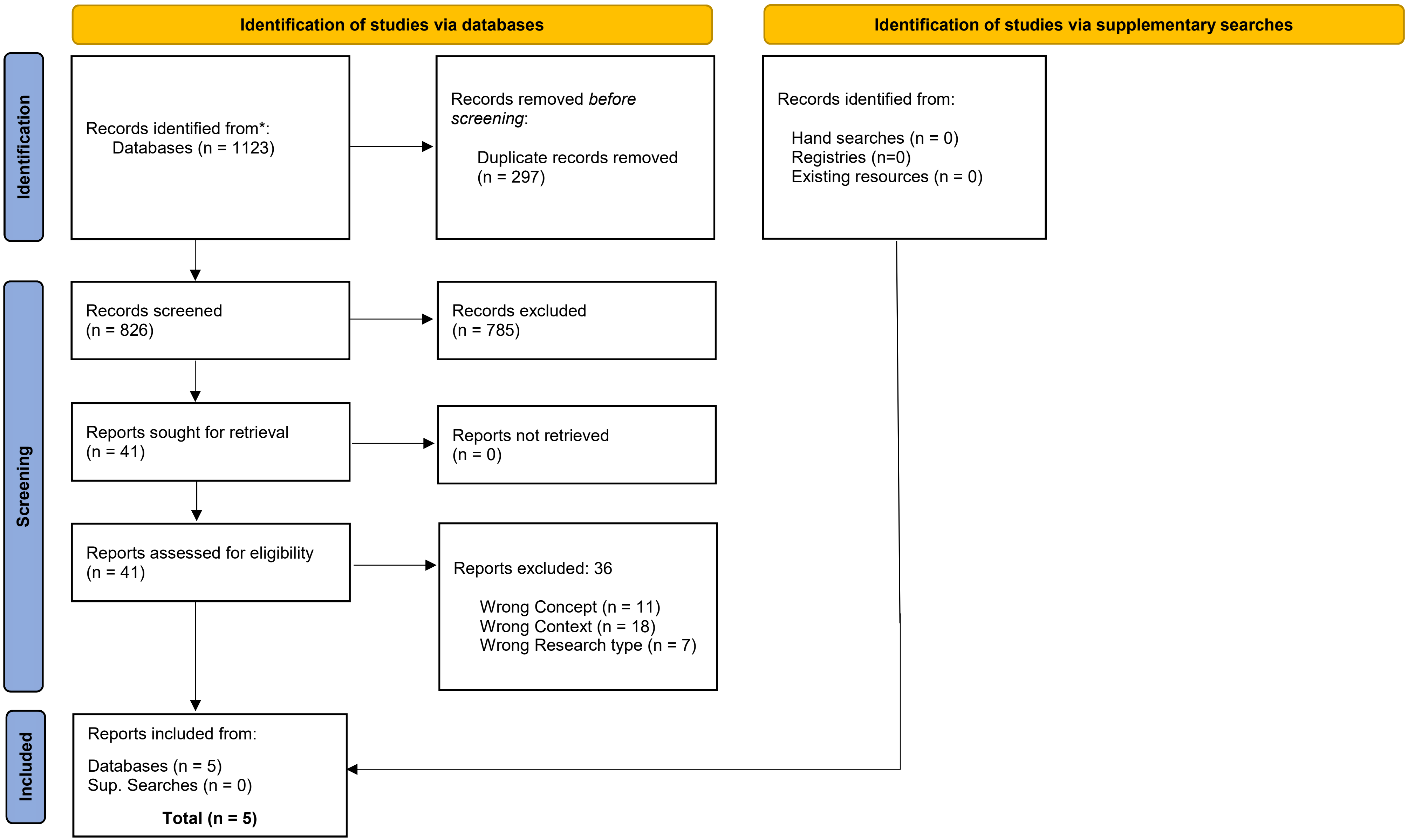


*Modified From:* Page MJ, McKenzie JE, Bossuyt PM, Boutron I, Hoffmann TC, Mulrow CD, et al. The PRISMA 2020 statement: an updated guideline for reporting systematic reviews. BMJ 2021;372:n71. doi: 10.1136/bmj.n71. For more information, visit: http://www.prisma-statement.org/
