## Supplemental File A4 for "Virtual Simulated Placements in Healthcare Education: A scoping review"

Supplementary Material A4: Revised data charting tool **(pilot revisions in bold)**

| DATA CHARTING | | | | |
|---|---|---|---|---|
| Author(s) | | | | |
| Year of publication | | | | |
| Title | | | | |
| Journal | | | | |
| Study location | | | | |
| **Pandemic response** | | | | |
| Study design **- comparator / control or pre-post measures** | | | | |
| Aims, objectives and research questions | | | | |
| Study population(s) | | | | |
| Sample size (total) | | | | |
| Number of groups | | Group 1 | Group 2 | Group 3 |
| Size of each Group | | | | |
| Intervention | **Scenario** | | | |
| Intervention description | **Activities** | | | |
| Intervention duration | | | | |
| **Intervention delivery** | **Software** | | | |
| **Intervention delivery** | **Hardware** | | | |
| **Stakeholders in the design** | | | | |
| Any underpinning concepts / theories **/ standards** | | | | |
| **Intended learning outcomes (ILOs) / capabilities** | | | | |
| Methodology | | | | |
| Student focussed outcome measures | | | | |
| Student outcomes | | | | |
| Important ~~results~~ **conclusions** | | | | |
| Sources of funding / conflicts of interest | | | | |
