## Supplemental File A5 for "Virtual Simulated Placements in Healthcare Education: A scoping review"

### Supplementary File A5: Table of included study characteristics

| Citation:<br><br>Author[1] (year)<br><br>Country[2]<br><br>Publication[3] | Purpose / aim | Population:<br><br>Profession (specialist rotation)[1]<br><br>Sample Size[2] | Study Design:<br><br>Comparison Groups[1]<br><br>Pre / post Measures[2] | VSP Design:<br><br>Stakeholders Involved[1]<br><br>Pedagogical Framework[2] | Desired capabilities | Methodology | Intervention:<br><br>Scenario(s)[1]<br><br>Activities[2]<br><br>Duration[3] | Delivery:<br><br>Software[1]<br><br>Hardware[2] | Student focussed outcome measures | Key Findings | Conclusions/ Implications | Funding / Conflicts of interest |
|---|---|---|---|---|---|---|---|---|---|---|---|---|
| [1] Alphert et al. (2021)[28]<br><br>[2] USA<br><br>[3] Academic Radiology | "To assess perceived student engagement & educational value of a new remote clinical radiology learning environment" p113 | [1] 2nd & 3rd year Medical students (on a Radiology rotation)<br><br>[2] 83 | [1] Conventional placement comparator group. Completed the in person course (mostly observing) within 6 months prior to the pandemic (n=36)<br><br>[2] post-test measures compared | [1] Not specified<br><br>[2] Not mentioned | Describing salient findings of images and working towards a diagnosis | Quantitative questionnaire | [1] Choice of 4 VRO specialties from: abdominal, breast, chest, emergency, musculoskeletal, neuroradiology & paediatric imaging<br><br>[2] VROs of curated cases<br>Online didactic & small group sessions.<br><br>[3] 4 weeks | [1] Webex, broadcasting Picture archiving and communicat-ion systems (PACS) workstations<br><br>[2] Screen based | Student experience:<br><br>Perceived sense of involvement, technical limitations & educational value of the learning experience | 87.2% & 75% response rates in grp 1 (VRO) & 2 (in person).<br><br>Educational value was comparable & interaction ratings were slightly higher in Grp 1: Perceived a more active role = 3.95(0.77), grp 2 =3.41(0.97) p=0.01. Reported less boredom = 1.93(0.47), grp 2 = 2.41(0.8) p=0.007<br><br>Confidence using the PACS was higher in grp 2 = 4.04(0.98), grp 1=2.3(1.16) p=0.0001 | Remote clinical radiology education can achieve a similar experience in fewer contact hours.<br><br>An advantage is the potential to standardise the education and provide a variety of images<br><br>A disadvantage is the lack of a full PACS workstation and socialisation with colleagues | Not Stated |

| Citation:<br><br>Author (year)[1]<br><br>Country[2]<br><br>Publication[3] | Purpose / aim | Population:<br><br>Profession (specialist rotation)[1]<br><br>Sample Size[2] | Study Design:<br><br>Comparison group[1]<br><br>pre / post measures[2] | VSP Design:<br><br>Stakeholders Involved[1]<br><br>Pedagogical framework[2] | Desired capabilities | Methodology | Intervention:<br><br>Scenario(s)[1]<br><br>Activities[2]<br><br>Duration[3] | Delivery:<br><br>Software[1]<br><br>Hardware[2] | Student focussed outcome measures | Key Findings | Conclusions/ Implications | Funding / Conflicts of interest |
|---|---|---|---|---|---|---|---|---|---|---|---|---|
| [1] Bhashyam and Dyer (2020)[29]<br><br>[2] USA<br><br>[3] Journal of the American Academy of Orthopaedic Surgeons | To create a learning platform to allow PGY-1 residents to develop basic orthopaedic knowledge & skills for emergency care & progress from basic to more advanced surgical procedures. | [1] Post graduate year one medical residents on an orthopaedic rotation<br><br>[2] 12 | [1] Single group<br><br>[2] Post-test design | [1] 1 x chief resident<br>1x program director<br>2x program coordinators<br><br>[2] Problem based learning was encouraged, given the evidence for benefit to adult learners | Cognitive knowledge and skill development | Quantitative questionnaire | [1] Basic skills/hand, sport, trauma & arthroplasty. 11 surgical techniques outlined<br><br>[2] Readings, videos, lectures & case "walk throughs." Videoconference demos & informal feedback on surgical skills<br><br>[3] 4 weeks | [1] Zoom<br><br>A central repository for pre-recorded lectures<br><br>[2] Screen based and use of home practice kits for surgical skills | Student satisfaction<br><br>Cost (total, start-up and recurring) | 100% were satisfied with the overall experience, module format, take home kits, helping their knowledge base & skill set. 92% felt it improved preparation for the operating room. Total cost per module per student is $1745 | Described a successful conversion of an in person placement to a virtual boot camp. This could be modified according to local policy, use with cadavers and VR when the technology becomes affordable | Not stated |
| [1] Creagh et al. (2021)[30]<br><br>[2] USA<br><br>[3] Clinical Imaging | “To meet the academic needs of medical students while providing a safe environment during the pandemic.<br><br>…To improve the residents' ability to teach.” p420 | [1] 3rd year Medical Students (on a radiology rotation)<br><br>[2] 41 | [1] Single group<br><br>[2] Post-test design | [1] Not specified<br><br>[2] Founded on the principles of andragogy:<br><br>Giving greater control over students’ own education.<br><br>Emphasis on problem-centred / experiential learning for relevance | Image interpretat-ion<br><br>Patient care & safety<br><br>Resource utilisation<br><br>Insight into the field<br><br>Differential diagnosis<br><br>Presentation and teaching skills | Quantitative methods to compare test scores and student evaluations | [1] Abdominal & spinal imaging. X-ray interpretation Women’s health Interventional radiography.<br><br>[2] Reading materials, lectures, modules American College of Radiography (ACR) e-learning<br>‘Hot seat’ sessions<br>OSCE style sessions<br>Conferences / tumour board meetings<br>Grand rounds presentations<br><br>[3] 4 weeks | [1] Aquifer (subscription based services)<br><br>Zoom<br><br>Webex<br><br>[2] Screen based | Student performance assessed with the AMSER (or ACR STARS) exam.<br><br>Evaluations on the content and structure. | Mean AMSER score was 75% (range 50–96%), matched the national average of 75%, $t(40) = -0.14868$, $p = .88$.<br><br>Positive feedback on the content, structure, engagement & time efficiency<br><br>Improvement in knowledge, leadership skills, search patterns & presenting findings | Student performance was in line with other courses on a national standardised examination.<br><br>The approach demonstrates the effectiveness of virtual radiology clerkships as viable alternatives to onsite rotations | Supported by HCA Healthcare<br><br>No conflicts of interest |

| Citation:<br><br>Author (year)[1]<br><br>Country<br><br>Publication[3] | Purpose / aim | Population:<br><br>Profession (specialist rotation)[1]<br><br>Sample Size[2] | Study Design:<br><br>Comparison group[1]<br><br>pre / post measures[2] | VSP Design:<br><br>Stakeholders Involved[1]<br><br>Pedagogical framework[2] | Desired capabilities | Methodology | Intervention:<br><br>Scenario(s)[1]<br><br>Activities[2]<br><br>Duration[3] | Delivery:<br><br>Software[1]<br><br>Hardware[2] | Student focussed outcome measures | Key Findings | Conclusions/ Implications | Funding / Conflicts of interest |
|---|---|---|---|---|---|---|---|---|---|---|---|---|
| [1] De Ponti et al. (2020)[31]<br><br>[2] Italy<br><br>[3]BMC Medical Education | "To assess medical students' perception on fully online training including simulated clinical scenarios during COVID-19 pandemic." p1 | [1] 6th year Medical students (Medicine & Surgery rotation)<br><br>[2] 122 | [1] Single group<br><br>[2] Post-test design | [1] Not specified<br><br>[2] Not mentioned | Clinical history taking,<br><br>Clinical decision making: ordering physical examination, laboratory /imaging tests and interventions | Quantitative questionnaire | [1] 21 simulated cases: 7 Cardiovascular & cerebrovascular cases<br>6 Trauma cases<br>2 Pneumonia cases<br>2 Infective disorder in pregnancy<br>2 Gastrointestinal surgery cases<br>1 Nephrological case<br>1 Hypoglycaemia case<br><br>[2] Introduction (to the case & software), virtual patient based training, debriefing<br><br>[3] 42 hours (21x 2hr) | [1] Body Interact<br><br>Microsoft Teams<br><br>[2] Screen based | Student satisfaction<br><br>Student feedback questionnaire | 115 (94%) response rate<br>90% gave a positive evaluation<br>93% appreciated the format<br>77% rated the VR realistic for the initial assessment, the diagnostic activity (94%) & treatment options (81%). 84% considered it useful for future hybrid training.<br>28% had technical issues with online access. | The online training avoided interruption to placements and the majority of participants gave a positive response (although a proportion reported technical difficulties) | None |
| [1] Durfee et al. (2020)[32]<br><br>[2] USA<br><br>[3] Academic Radiology | "Describe the design and the logistical challenges involved in structuring a virtual radiology clerkship and assess its efficacy." p1462 | [1] Medical students in a radiology rotation<br><br>[2] 111 | [1] Single group<br><br>[2] Post-test design | [1] The clerkship directors from three hospitals<br><br>[2] Not mentioned | Patient and safety centred focus Image utilisation, interpretatio n and generation of a differential diagnoses | Quantitative student scores and questionnaires | [1] 19 Aquifer modules (no detail of the cases provided)<br><br>[2] Large group didactic lectures<br>Small group homeroom activities: Topic of the day (flipped classroom) and an unknown case conference (readout session) | [1] Aquifer<br><br>Zoom<br><br>Poll Everywhere<br><br>[2] Screen based | Student performance on the AMSER exam<br><br>Student feedback | AMSER scores averaged 85% (64-95%): comparable to the in person course.<br><br>50% response rate: 100% rated the course overall as good/excellent.<br><br>Suggested improvements commonly related to the didactic lectures | The virtual radiology core clerkship was a successful educational experience for medical students. Students enjoyed the small group homerooms, although personal connections were challenging | None stated |

| Citation: Author (year)[1] Country[2] Publication[3] | Purpose / aim | Population: Profession (specialist rotation)[1] Sample Size[2] | Study Design Comparison group[1] pre / post measures[2] | VSP Design: Stakeholders Involved[1] Pedagogical framework[2] | Desired capabilities | Methodology | Intervention: Scenario(s)[1] Activities[2] Duration[3] | Delivery: Software[1] Hardware[2] | Student focussed outcome measures | Key Findings | Conclusions/ Implications | Funding / Conflicts of interest |
|---|---|---|---|---|---|---|---|---|---|---|---|---|
| [1] Fehl et al. (2022)[33]<br>[2] Germany<br>[3] Medical Education Online | To provide students an insight into general practice with its particularities regarding patient clientele, spatial conditions and economic and organisational structure despite the lack of physical presence" p2 | [1] 4th year Medical students (in GP practice)<br>[2] 192 | [1] Conventional clerkship group (n=277)<br>[2] Post-test comparison of student evaluations | [1] Not specified<br>[2] Principles of 'good online teaching' i.e. clear learning objectives matching the curriculum, synchronous /asynchronous teacher-student interaction, promotion of higher-order thinking & communication skills, encouragement of active & self-directed learning while promoting timely completion of tasks & effective time management | Higher-order thinking<br>Communicat-ion skills | Mixed: Surveys generated quantitative & qualitative data which were analysed separately | [1]SOAP cases: gout, acute vertigo, sore throat, hypertension check-up, acute burning on urination, subacute chest pain, geriatric home visit meds review, vaccination, prolonged cough & acute back pain<br>[2] 10 SOAP cases partly linked with physical examination videos<br>Videos / materials to learn about general practice.<br>Visual diagnosis from images<br>Live video chats with GP teachers<br>[3] 2 weeks | [1] Student portal<br>Email<br>Video chat<br>Telephone calls (optional)<br>[2] Screen based Phone | Working enjoyment<br>Learning gain<br>Practical relevance Insight into GP work<br>Usage behaviours (devices & chosen teaching formats).<br>Open questions: 'What did they like about the virtual clerkship?' 'What could be improved?' | 51.6% response rate for group 1 & 100% for group 2. Group 1: 87.9% enjoyed it 89.9% learned a lot, 76.8% gained practical insights 90.9% perceived high practical relevance. 65.6% welcomed this format into future clerkships.<br>89% laptop usage. Clinical cases, videos, visual diagnosis and communication with the GP teachers were valued the most. Students recommended an increase in clinical case content.<br>Comparison: The acquisition of new skills & attitudes were rated superior in the conventional clerkship. | Students welcomed the digital clerkship.<br>The flexible time management, structure & multifaceted learning content were valued.<br>It rated comparably to face to face (FTF) learning overall, but online was considered better for teaching theoretical rather than practical skills<br>FTF GP clerkships may benefit from complementing online teaching, in a blended approach. | None |

| Citation:<br><br>Author (year)[1]<br><br>Country[2]<br><br>Publication[3] | Purpose / aim | Population:<br><br>Profession (specialist rotation)[1]<br><br>Sample Size[2] | Study Design:<br><br>Comparison group[1]<br><br>pre / post measures[2] | VSP Design:<br><br>Stakeholders Included[1]<br><br>Pedagogical framework[2] | Desired capabilities | Methodology | Intervention:<br><br>Scenario(s)[1]<br><br>Activities[2]<br><br>Duration[3] | Delivery:<br><br>Software[1]<br><br>Hardware[2] | Student focussed outcome measures | Key Findings | Conclusions/ Implications | Funding / Conflicts of interest |
|---|---|---|---|---|---|---|---|---|---|---|---|---|
| [1] Ganji et al. (2022)[34]<br><br>[2] Iran<br><br>[3] Nurse Education Today | To determine the effect of a virtual gynaecology clinic training programme on the knowledge and clinical skills of midwifery students | [1] Midwifery interns on a Gynaecology rotation<br><br>[2] 47 | [1] Single group<br><br>[2] Repeated measures design | [1] Research team and midwifery experts (faculty members & senior lecturers). Students were consulted via an educational needs interview<br><br>[2] ADDIE model: (Analysis Design Development Implementation Evaluation) | Knowledge<br><br>Clinical skills in Interview & history taking<br><br>Problem evaluation<br><br>Clinical judgement<br><br>Problem assessment/ management | Reports on the quantitative part of a mixed methods study | [1] 27 cases including genital infections, abnormal bleeding, menopause, ovarian cysts & abnormal smears<br><br>[2] Multiple choice Webinars, Videos Cases with questions regarding interview, diagnoses & treatment.<br><br>[3] Not stated but 2 days were allocated for each of the 4 stages of the cases | [1] Navid – Learning management system (LMS)<br><br>WhatsApp<br><br>Adobe connect<br><br>[2] Screen based | Knowledge<br><br>Skills: Modified Mini- CEX - rated over 4 virtual cases.<br><br>Student satisfaction on a scale of 1-9 | Knowledge scores pre & post learning were $10.0 \pm 1.74$ & $13.80 \pm 1.43$, $p < 0.001$.<br><br>Post-training scores improved from satisfactory to excellent for clinical judgment, consultation efficiency & clinical competence. Interview scores increased but remained in the satisfactory range. | Training through virtual clinic promoted the knowledge & clinical skills of midwifery interns.<br><br>A virtual clinic may be used in crisis situations & in combination with teaching under normal circumstances by strengthening the infrastructure & removing barriers. | Financially supported by the university.<br><br>No conflict of interest |
| [1] Gomez et al (2021)[35]<br><br>[2] USA<br><br>[3] Academic Radiology | To rapidly convert to an in-person diagnostic radiology elective to a remote learning experience. | [1] 2nd, 3rd & 4th year Medical students (Radiology rotation)<br><br>[2] 116 | [1] Single group<br><br>[2] Post-test design | [1] Course directors<br><br>[2] Not mentioned | Knowledge<br><br>Skills in identifying normal anatomy and common pathology | Mixed data in the survey responses | [1] Not specified<br><br>[2] Interactive cases. Readouts & Hot seat cases Quizzes & jeopardy Website resources Zoom chats Q&A Narrated PowerPoint student submissions Journal club discussion<br><br>[3] 3 weeks | [1] Pacsbin (image library) Education websites & modules Zoom Nearpod PowerPoint Microsoft & Google forms Microsoft Teams Blackboard<br>[2] Screen based | Learner achievement (via completion of quizzes)<br><br>Final exam (modified to reflect the altered course content)<br><br>Enrolment metrics<br><br>Student feedback | 100% pass rate<br><br>Largely positive feedback & gratitude for the opportunity to continue learning<br><br>Recommended more, small group learning, interactive / reflective content, trainee led teaching & shorter days | The current state of technology makes radiology particularly well suited for distance learning, & with the proper tools and approaches, effective remote radiology instruction can be achieved. | None mentioned |

| Citation:<br><br>Author (year)[1]<br><br>Country[2]<br><br>Publication[3] | Purpose / aim | Population:<br><br>Profession (specialist rotation)[1]<br><br>Sample Size[2] | Study Design:<br><br>Comparison group[1]<br><br>pre / post measures[2] | VSP Design:<br><br>Stakeholders Involved[1]<br><br>Pedagogical framework[2] | Desired capabilities | Methodology | Intervention:<br><br>Scenario(s)[1]<br><br>Activities[2]<br><br>Duration[3] | Delivery:<br><br>Software[1]<br><br>Hardware[2] | Student focussed outcome measures | Key Findings | Conclusions/ Implications | Funding / Conflicts of interest |
|---|---|---|---|---|---|---|---|---|---|---|---|---|
| [1] He et al. (2021)[36]<br><br>[2] China<br><br>[3] Medical education online | To examine the effect of an online neurology course & whether it can cater for interns from different programs.<br><br>Whether group size has an impact. To analyse how it can be refined. | [1] Medical interns (on a neurology rotation)<br><br>[2] 92 | [1] Single group<br><br>[2] Post-test design with subgroup analysis by:<br><br>Programme enrolment (3 groups)<br><br>Intake (6 groups) | [1] Not specified<br><br>[2] Not mentioned | Practical skills (New patient admission, physical exam & medical record writing).<br><br>Theoretical knowledge | Quantitative | [1] Nervous system, Cardiopulmonary resuscitation & Lumbar puncture<br><br>[2] Small private online courses (SPOC). Didactic, flipped classroom & case based learning. Videos of ward rounds, typical clinical cases & difficult case discussions. Interactive case discussions /conferences & reading<br><br>[3] 3 weeks | [1] Tencent class (live broadcast platform).<br><br>WeChat group (for shared files)<br><br>PowerPoint<br><br>[2] Screen based | Final exam scores<br><br>Student evaluation | 100% response rate & consistent positive ratings<br><br>99% recommended incorporating the course into future programmes.<br><br>No difference in test scores between programs ($p < 0.05$)<br><br>Students groups < 15 had a better learning experience ($p < 0.05$)<br><br>Interactive discussions & analysis were rated highest. | The neurology training course was effective and was highly rated by the interns. | Funding from: Central South University. National Natural Science Foundation of China. Huxiang High-Level Talent Gathering Project.<br><br>No conflicts of interest |
| [1] Holmberg et al. (2021)[37]<br><br>[2] USA<br><br>[3] Academic Medicine | To deliver essential elements of the sub-internship virtually and to address limited teaching faculty availability | [1] 4th year Medical students on an internal medicine sub-internship<br><br>[2] 10 | [1] Single group<br><br>[2] Repeated measures design | [1] Clerkship directors, course director, recent graduates of the in-person course (4 senior medical students - near peers)<br><br>[2] Not mentioned | Order-writing Communication<br><br>Clinical reasoning<br><br>Using medical literature<br><br>Admitting a patient, cross overs & handoffs<br><br>Independent learning | Mixed data in the survey responses | [1] Not specified<br><br>[2] Orientation & debrief. Student presentations Interactive lectures Small-group discussions Case-based faculty led/peer teaching Role-play Resident report<br><br>[3] 4 weeks | [1] Zoom<br><br>[2] Screen based | Student experience<br><br>5 self-rated competencies<br><br>The extent to which the course accomplished its learning objectives | All self-rated competencies demonstrated significant improvement except the describing how to efficiently admit a patient (didn't reach statistical significance).<br><br>Open-ended responses indicated initial skepticism, but the course exceeded expectations. | Our findings and our experiences with the virtual sub-internship suggest that a virtual sub-internship can be a high quality educational experience | None |

| Citation:<br><br>Author (year)[1]<br><br>Country[2]<br><br>Publication[3] | Purpose / aim | Population:<br><br>Profession (specialist rotation)[1]<br><br>Sample Size[2] | Study Design:<br><br>Comparison groups[1]<br><br>pre / post measures[2] | VSP Design:<br><br>Stakeholders Involved[1]<br><br>Pedagogical Framework[2] | Desired capabilities | Methodology | Intervention:<br><br>Scenario(s)[1]<br><br>Activities[2]<br><br>Duration[3] | Delivery:<br><br>Software[1]<br><br>Hardware[2] | Student focussed outcome measures | Key Findings | Conclusions/ Implications | Funding / Conflicts of interest |
|---|---|---|---|---|---|---|---|---|---|---|---|---|
| [1] Joung and Kang (2022)[38]<br><br>[2] South Korea<br><br>[3] Issues in Mental Health Nursing | Investigate the potential of VS-based education as an alternative for clinical psychiatric nurse training & consider how it can be optimised as an educational method | [1] 4th year Nursing Students (Psychiatry rotation)<br><br>[2] 20 | [1] Single group<br><br>[2] Post-test design | [1] Not specified<br><br>[2] Not mentioned | The transfer of intrinsic nursing values such as empathy. | Qualitative | [1] Schizophrenia, bipolar disorder, anxiety disorder and depressive disorder<br><br>[2] vSim sessions, team conferences with an instructor<br><br>[3] 90 hours over 10 days | [1] vSim for Nursing Video conferencing software<br><br>[2] Screen based | Focus Groups | 3 key themes:<br>1. Students were glad that the patients were not real people<br>2. vSim serving as a bridge between the text & real world<br>3. Supplementat-ions needed for vSims to replace clinical practice | vSim was recognised as a tool linking theory with actual clinical practical training. Students were able to repeat practice to solve problems and work in a safe environment but were unable to have real human experiences. | No funding or conflict of interest |
| [1] Kasai et al. (2021)[39]<br><br>[2] Japan<br><br>[3] BMC Medical Education | To evaluate the feasibility & effectiveness of this approach. To identify the advantages & disadvantage s of online-simulated clinical placement (sCP) from the medical students' perspectives | [1] 5th year Medical students on a respiratory unit & general medicine rotation<br><br>[2] 43 | [1] Single group<br><br>[2] Repeated measures design | [1] Not specified<br><br>[2] Peer assisted learning applied to problem based learning | History taking<br><br>Diagnosis<br><br>Select tests & interpret results<br><br>Treatment planning<br><br>Medical documentation & present the clinical course<br><br>Perform safe, evidence based treatment<br><br>Informed consent & patient education | Mixed | [1] General medicine outpatient cases & respiratory inpatient cases<br><br>[2] Simulated electronic & health records (sEHR) electronic-Practice Based Learning (e-PBL) Online virtual medical interviews (VMI)<br><br>[3] 4 weeks | [1] Video conference system<br><br>Learning management system (LMS)<br><br>Microsoft excel<br><br>[2] Screen based | Self-evaluation of clinical performance<br><br>Semi structured focus groups | 100% response rate<br><br>Online sCP is an efficient learning method & useful for learning how to write medical records & summaries<br><br>Clinical clerkship was more useful for learning associated with medical interviews, physical examination & humanistic qualities eg. professionalism | Online-sCP with sEHR, e-PBL, and online-VMI could be useful in learning some of the clinical skills acquired through clinical clerkship. | None |

| Citation<br><br>Author (year)[1]<br><br>Country[2]<br><br>Publication[3] | Purpose / aim | Population:<br><br>Profession (specialist rotation)[1]<br><br>Sample Size[2] | Study Design:<br><br>Comparison group[1]<br><br>pre / post measures[2] | VSP Design:<br><br>Stakeholders Involved[1]<br><br>Pedagogical Frameworks[2] | Desired capabilities | Methodology | Intervention:<br><br>Scenario(s)[1]<br><br>Activities[2]<br><br>Duration[3] | Delivery:<br><br>Software[1]<br><br>Hardware[2] | Student focussed outcome measures | Key Findings | Conclusions/ Implications | Funding / Conflicts of interest |
|---|---|---|---|---|---|---|---|---|---|---|---|---|
| [1] Kubin et al. (2021)[40]<br><br>[2] USA<br><br>[3] Journal of Nursing Education | To develop an innovative re vised plan for facilitation of clinical learning experiences in the virtual learning environment. | [1] Nursing students on a Paediatric Rotation<br><br>[2] Not stated (taught in small groups of 5-10) | [1] Single group<br><br>[2] Post-test design | [1] Not specified<br><br>[2] International Nursing Association of Clinical Simulation and Learning (INACSL) best practice guidelines | Nursing process<br><br>Growth and development<br><br>Assessment<br><br>Clinical judgment & reasoning skills<br><br>Prioritisation & delegation<br><br>Communicati on skills. | Mixed: Survey with Likert & open ended questions | [1] Various paediatric disorders. Child with diabetic ketoacidosis was used in the escape room<br><br>[2] Virtual escape rooms, unfolding video case studies, & blended prioritisation simulations.<br><br>[3] Not stated | [1] vSim<br><br>NurseThink vClinical<br><br>F.A. Davis' Paediatric Interactive Clinical Scenarios<br><br>Virtual Healthcare Experience<br><br>Flipgrid<br><br>Google Forms & Sites<br><br>[2] Screen based | Student satisfaction.<br><br>Evaluations pf each clinical activity and the ability to meet course outcomes | 100% response rate<br><br>Self-reported increases in clinical reasoning, prioritisation, communication and critical thinking skills. | Virtual activities can be as effective as in-person clinical learning methodologies. Integrating virtual activities into clinical curricula can be a viable option, especially in areas where clinical placement is limited | Not stated |
| [1] Luo et al. (2021)[41]<br><br>[2] China<br><br>[3] Clinical Simulation in Nursing | To understand students' performance, learning effectiveness & satisfaction with their participation in distance learning.<br>To compare outcomes between genders | [1] 4th year Nursing students<br><br>[2] 35 | [1] Single group<br><br>[2] Repeated measures design | [1] Nursing educators from the University<br><br>[2] Outcome-Based Educational theory, which emphasises student centred learning<br><br>National Standards for Nursing Undergraduates | Pass the 2020 Chinese Registered Nurse Licensure Exam<br><br>knowledge and clinical competence requirements specified in the National Standards | Quantitative | [1] Medical, surgical, obstetrics & gynaecology, paediatrics, fundamental nursing<br><br>[2] Webinars (lectures & case based learning) & Virtual simulations<br><br>[3] 3 months | [1]Videoconfere ncing platforms (Tencent Meeting & Ding Talk)<br><br>vSim<br><br>[2] Screen based | Theoretical knowledge<br><br>Clinical thinking ability<br><br>Academic self-efficacy<br><br>Student satisfaction | 100% response rate<br><br>High levels of student engagement, satisfaction & theoretical knowledge. Significant improvements in Systematic, Evidence based & Clinical thinking Females outperformed males in all domains | Distance learning combining webinars & virtual simulations could meet the learning requirements of senior nursing students in a safe environment in a flexible manner, & students could obtain theoretical knowledge & grow their clinical thinking ability | Funded by Wuhan University Teaching & Research reform Project.<br><br>No conflicts of interest |

| Citation:<br><br>Author (year)[1]<br><br>Country[2]<br><br>Publication[3] | Purpose / aim | Population:<br><br>Profession (specialist rotation)[1]<br><br>Sample Size[2] | Study Design:<br><br>Comparison groups[1]<br><br>pre / post measures[2] | VSP Design:<br><br>Stakeholders Involved[1]<br><br>Pedagogical Framework[2] | Desired capabilities | Methodology | Intervention:<br><br>Scenario(s)[1]<br><br>Activities[2]<br><br>Duration[3] | Delivery:<br><br>Software[1]<br><br>Hardware[2] | Student focussed outcome measures | Key Findings | Conclusions/ Implications | Funding / Conflicts of interest |
|---|---|---|---|---|---|---|---|---|---|---|---|---|
| [1] Martin-Delgado et al (2022)[42]<br><br>[2] Spain<br><br>[3] Journal of Professional Nursing | To explore final-year nursing experiences from completing their clinical training in a teaching role practicum during the pandemic. | [1] Final year Nursing students<br><br>[2] 34 | [1] Single group<br><br>[2] Post-test design | [1] Not specified<br><br>[2] Not stated | Designing and developing evidence based educational materials aimed at meeting the learning needs of their peers. | Qualitative | [1] Covid 19 educational needs, including management of respiratory patients, mechanical ventilation, use of protection equipment<br><br>[2] Online training Mentoring sessions Design and development of educational material<br><br>[3] 3 months | [1] Video conferencing software, Moodle (LMS)<br><br>[2] Screen based | Themes from student reflective journals (18 of the 34 students) | Three themes<br>1. Emotions due to not being able to complete their final placement & not to joining the workforce<br>2. Perceived benefits of a teaching role,<br>3. Recognising the teaching role as key to the profession & the importance of scientific evidence in clinical practice. | The online teaching practicum gave students the opportunity to develop education competencies. | No mention of conflicts of interest. No external funding |
| [1] Nguyen et al (2023)[43]<br><br>[2] USA<br><br>[3] MedEdPortal | To transition an intro-ductory anesthesi-ology clerkship to an entirely virtual curriculum | [1] 3rd & 4th year Medical students (anaesthesi-ology)<br><br>[2] 28 | [1] Single group<br><br>[2] Post-test design | [1] Not stated, but past student surveys from placements were used for a needs assessment<br><br>[2] Kerns 6 steps of curriculum development | Nine educational objectives were outlined, including information/ description, differential diagnosis, treatment prioritisation and planning | Mixed | [1] Preoperative evaluation, inhaled intravenous anaesthetics, airway management, anaphylaxis, malignant hyperthermia and unanticipated difficult airway<br><br>[2] Didactics, assigned readings, case based learning discussions<br><br>[3] 2 weeks | [1] Canvas (LMS) PowerPoint Simulation videos Zoom<br><br>[2] Screen based | Survey with Likert and open-text responses | 79% response rate. Clerkship met / exceeded expectations in all areas. All students agreed / strongly agreed that the objectives were clear & achieved. Two students indicated that the assessment tools could align better with the curriculum & one wanted more didactics. One noted technical issues with Zoom | A compelling clerkship was executed, which was highly rated. | No disclosures or funding to report |

| Citation<br><br>Author (year)[1]<br><br>Country[2]<br><br>Publication[3] | Purpose / aim | Population:<br><br>Profession (specialist rotation)[1]<br><br>Sample Size[2] | Study Design:<br><br>Comparison group[1]<br><br>pre / post measures[2] | VSP Design:<br><br>Stakeholders Involved[1]<br><br>Pedagogical framework[2] | Desired capabilities | Methodology | Intervention:<br><br>Scenario(s)[1]<br><br>Activities[2]<br><br>Duration[3] | Delivery:<br><br>Software[1]<br><br>Hardware[2] | Student focussed outcome measures | Key Findings | Conclusions/ Implications | Funding / Conflicts of interest |
|---|---|---|---|---|---|---|---|---|---|---|---|---|
| [1] Rahm et al. (2021)[44]<br><br>[2] Germany<br><br>[3] PLOS One | To enrich our understanding of how students perceive realistic multimodal game-like e-learning cases within a complete e-learning-based curriculum. | [1] Medical students in an internal medicine rotation<br><br>[2] 198 | [1] Single group<br><br>[2] Post-test design | [1] Medical students (who had already completed the internal medicine module) and physicians from different disciplines<br><br>[2] Not stated | Decision-making skills<br><br>Communication<br><br>Diagnostic thinking | Mixed: Quantitative survey with free text space for student feedback | [1] Cases based on routine encounters across different clinical settings<br><br>[2] e-learning cases with quizzes & interaction modules with gamification<br><br>[3] 10 weeks | [1] articulate.com (bespoke creator tool)<br><br>Moodle LMS<br><br>[2] Screen based | Student evaluation | 49.5 to 82.5% response rates to case evaluations & 25.8% end of term response<br><br>Clinical context, interactivity, game-like interface & embedded learning in the cases motivated students to engage with the learning materials & cases | Solving and interpreting e-learning cases close to real-life settings promoted students' motivation during the COVID-19 pandemic and may partially have compensated for missing bedside teaching opportunities. | Funded by clinician-scientist-program of the German Internal Medicine Society (DGIM).<br><br>No conflicts of interest |
| [1] Redinger and Greene (2021)[45]<br><br>[2] USA<br><br>[3] Western Journal of Emergency Medicine | To describe the development, application & program evaluation of a virtual advanced emergency medicine (EM) curriculum developed rapidly in response to the COVID-19 pandemic. | [1] 4th year Medical students on an EM rotation<br><br>[2] 104 | [1] Traditional rotation from a previous cohort (n= 48)<br><br>[2] Post-test exam scores compared between groups | [1] Not Specified<br><br>[2] Kerns 6 step model for curriculum development | History, physical, diagnosis & case presentation<br><br>Common diagnostic studies,<br><br>Management plans,<br><br>Knowledge, indications / constraints & basic procedural skills.<br><br>Emergency recognition & management | Mixed methods | [1] 12 most common chief complaints in clinic<br><br>[2] Case series, radiology & ECG interpretation, textbooks, journal articles, podcasts, online board review, blog posts quizzes & a case presentation.<br><br>[3] 4 weeks | [1] Microsoft Teams<br><br>MedEd Case X<br><br>EM: RAP C3 series<br><br>SAEM EM Curriculum<br><br>Sublux Radiology App<br><br>A Night in the ER App<br><br>[2] Screen based | Student performance (National Standardised EM Shelf Exam) – simplified to pass/fail<br><br>Course evaluation (focus group) | No difference between performance scores $t(102) = 1.317$ $p = 0.174$<br><br>Comments indicate that the virtual clerkship successfully met their learning needs, resulting from its design, organisation & use of quality learning resources. | Students demonstrated the same levels of knowledge in the virtual & traditional rotations<br><br>Feedback was overall positive, although limited peer interaction & group learning dynamics were noted. | None |

| Citation:<br><br>Author (year)¹<br><br>Country²<br><br>Publication³ | Purpose / aim | Population:<br><br>Profession (specialist rotation)¹<br><br>Sample Size² | Study Design:<br><br>Comparison groups¹<br><br>pre / post measures² | VSP Design<br><br>Stakeholders Involved¹<br><br>Pedagogical Frameworks² | Desired capabilities | Methodology | Intervention:<br><br>Scenario(s)¹<br><br>Activities²<br><br>Duration³ | Delivery:<br><br>Software¹<br><br>Hardware² | Student focussed outcome measures | Key Findings | Conclusions/ Implications | Funding / Conflicts of interest |
|---|---|---|---|---|---|---|---|---|---|---|---|---|
| [1] Samueli et al. (2020)[46]<br><br>[2] Israel<br><br>[3] Annals of Diagnostic Pathology | To review a diagnostic pathology selective for undergrad medical Students. Including the design, operation, evaluation, & suggestions for further adjustments. | [1] 3rd and 4th year medical students (diagnostic pathology selective)<br><br>[2] 59 | [1] Single group<br><br>[2] Post-test measures | [1] Course coordinator<br><br>[2] Kerns 6 step framework for curriculum development | Introduce surgical pathology<br><br>Reinforce the pathological basis for disease, including mechanisms & treatments<br><br>Appreciate "the way a pathologist thinks," & what they "mean" in their reports, as well as the significance of commonly describ ed findings | Quantitative Survey (with open text options) | [1] Principles of non-neoplastic (inflammatory) & neoplastic (benign/malignant) disorders, Dermatopathology, Breast pathology, Neoplastic neuropathology, Neoplastic thyroid pathology Advanced topics in diagnostic pathology (NUT carcinoma, thyroid pathology)<br><br>[2] Self-assigned reading , lectures, slide reviews, diagnostic quiz<br><br>[3] 2 weeks | [1] Zoom PowerPoint Moodle<br><br>Whole slide image (WSI) viewers: (CaseViewer & Aperio ImageScope)<br><br>Library subscription (for assigned texts)<br><br>[2] Screen based | Student experience (previous exposure to pathology), Level of interest & learning from the course, Evaluation / feedback | 42% survey response rate, Participants new to diagnostic pathology instruction. Overall, the course was rated very favourably: 68% gave at least 3 out of 4 points for questions related to course interest, improved understanding of diseases & how strongly they would recommend the course. The key disadvantage as reported by 80% was tech issues accessing the slides | The course was a success and can be a model for future virtual pathology electives.<br><br>Great effort should made to provide technical support to the students.<br><br>The selective demonstrated value for students and provided much-needed exposure to diagnostic pathology in clinical practice. | None |

| Citation:<br><br>Author (year)[1]<br><br>Country[2]<br><br>Publication[3] | Purpose / aim | Population:<br><br>Profession (specialist rotation)[1]<br><br>Sample Size[2] | Study Design:<br><br>Comparison groups[1]<br><br>pre / post measures[2] | VSP Design:<br><br>Stakeholders Involved[1]<br><br>Pedagogical Framework[2] | Desired capabilities | Methodology | Intervention:<br><br>Scenario(s)[1]<br><br>Activities[2]<br><br>Duration[3] | Delivery:<br><br>Software[1]<br><br>Hardware[2] | Student focussed outcome measures | Key Findings | Conclusions/ Implications | Funding / Conflicts of interest |
|---|---|---|---|---|---|---|---|---|---|---|---|---|
| [1] Smith and Jones (2023)[47]<br><br>[2] Australia<br><br>[3] BMC Medical Education | Provide an elective that enables students to make the current clinical world relevant, cover key content to assure intern preparedness & explore how COVID-19 changed one key area of medical practice. | [1] 4th year Medical students<br><br>[2] 250 | [1] Single group<br><br>[2] Post-test design | [1] Academics and professional support (clinicians)<br><br>[2] Not mentioned | Clinical communication<br><br>To author case studies of COVID approaches | Mixed | [1] Atrial fibrillation, depression, hypertension, prone ventilation, using protective equipment, lung ultrasound. COVID Global health, public health, child health, aged care, legal & ethical, general practice. Primary care, mental health & evidence-based practice.<br><br>[2] Podcasts, case studies, flipped classrooms, tutorials & modules<br><br>[3] 6 weeks (200 hrs) | [1] Microsoft Teams PowerPoint OSLER (logging of progress & assessment) National Prescribing Service (NPS) modules COVID online modules.<br><br>[2] Screen based | Evaluation survey with Likert responses and open-ended questions | 32% response rate. Overall worked well, was well coordinated & a good option for a disrupted placement. The project options met their needs very well & were well supervised. More guidance asked for on COVID information & academic writing support for publication. Some found the OSLER and NPS modules a bit dry. | The COVID-19 e-elective was successful in meeting student learning needs & alleviated the concerns of students whose placements were disrupted. | No external funding and no competing interests |
| [1] Steehler et al. (2021)[48]<br><br>[2] USA<br><br>[3] Otolaryngology - Head and Neck Surgery | To develop and evaluate a virtual otolaryngology elective created during COVID-19.<br><br>To teach the basics of otolaryngology & increase exposure to the specialty | [1] 3rd & 4th Medical students (head & neck surgery rotation)<br><br>[2] 12 | [1] Single group<br><br>[2] Repeated measures design (for n=5 pre & post knowledge test scores) | [1] Faculty, residents and senior medical students<br><br>[2] Not mentioned | Pathophys-iology<br><br>Workup<br><br>Differential diagnosis<br><br>Treatment of disease course<br><br>otolaryngology practice & referral | Mixed methods | [1] Rhinology, otology, facial plastic & reconstructive, laryngology, paediatric otolaryngology, imaging, & emergencies.<br><br>[2] Orientation, anatomy/examination & surgical videos, reading, lectures, case based learning, grand rounds & roundtable conversation<br><br>[3] 1 week | [1] Zoom<br><br>[2] Screen based | Test scores (pre & post for n=5)<br><br>Student evaluation | 92% reported increased understanding & interest in the field<br><br>Increase in knowledge test scores (p=0.001).<br><br>Appreciation for course organisation, formative assessment & case based learning | An virtual otolaryngology elective format can be effective at providing an educational experience & garnering interest | None |

| Citation:<br>Author (year)[1]<br>Country[2]<br>Publication[3] | Purpose / aim | Population:<br>Profession (specialist rotation)[1]<br>Sample Size[2] | Study Design:<br>Comparison group[1]<br>pre / post measures[2] | VSP Design:<br>Stakeholders Involved[1]<br>Pedagogical frameworks[2] | Desired capabilities | Methodology | Intervention:<br>Scenario(s)[1]<br>Activities[2]<br>Duration[3] | Delivery:<br>Software[1]<br>Hardware[2] | Student focussed outcome measures | Key Findings | Conclusions/ Implications | Funding / Conflicts of interest |
|---|---|---|---|---|---|---|---|---|---|---|---|---|
| [1] Taylor et al. (2021)[49]<br>[2] UK<br>[3] British Journal of Nursing | To explore & discuss a simulated clinical placement, aimed at enhancing the learning experience to create effective, efficient clinicians | [1] 2st year Dietetics students<br>[2] 40 | [1] Single group<br>[2] Post-test design | [1] Not specified<br>[2] Controlled reflective processes underpinned the online workbook<br>NHS & HCPC placement standards | Knowledge, communication & professional practice domains | Mixed methods | [1] Not specified, but simulated patient journeys<br>[2] Statutory & mandatory training Virtual wards & mealtimes. Recordings, peer learning with structured activities, & an online workbook<br>[3] 2 weeks | [1] 360 images<br>Diet-COMMS<br>COVCollaborate App<br>Microsoft Teams<br>[2] Screen based | Student evaluation (questionnaire & focus group)<br>Web page metrics<br>Student results | 100% of the cohort passed the placement.<br>360 images: Rated enjoyable & informative. Were accessed 1016 times | Despite some concerns / issues, a virtual placement can be a useful, rich experience for the student. | Funding not mentioned. No conflicts of interest |
| [1] Villa et al. (2021)[50]<br>[2] USA<br>[3] Western Journal of Emergency Medicine | To create, implement & evaluate a virtual clerkship with a focus on social emergency medicine (EM) & profession-al develop-ment | [1] 4th year Medical students on an EM Clerkship<br>[2] 26 | [1] Single group<br>[2] Repeated measures design | [1] Clerkship director, associate programme directors, medical education fellows and senior EM residents. Needs assessment of post graduate near peers.<br>[2] Kern's method for curriculum development<br>Framework for maximising online learning | Advanced medical Knowledge<br>Social determinants of health<br>Professional development<br>Professional identity formation. | Mixed methods | [1] Paediatric anaphylaxis, motorcycle trauma, hypothermia & abdominal aortic aneurysm Themes: language, incarceration, gender identity, race & homelessness<br>[2] Assignments (using websites & podcasts), small group didactic sessions, student led teaching, virtual escape rooms & book club.<br>[3] 2 weeks | [1] Zoom<br>Foundations of EM (online resource)<br>IDHEAL modules<br>[2] Screen based | Pre & post knowledge tests<br>Survey evaluations:<br>Overall attitude to the course<br>After each module x 5 (to determine the comfort with applying content to a clinical setting) | 75% & 96% survey response rates: post-module & end-rotation<br>Modest gains in knowledge scores ($p=0.006$, effect size: 0.68, 95% CI 0.12-1.24)<br>89% strongly agreed: topics were important.<br>95% strongly agreed /agreed: rotation should be repeated<br>Positive feedback for course design, but zoom fatigue was mentioned. | A virtual EM visiting clerkship is feasible, supports knowledge acquisition & is perceived as valuable by participants.<br>Virtual learning experiences may be valuable in the future as an adjunct to traditional in-person rotations. | None |

| Citation:<br><br>Author (year)[1]<br><br>Country[2]<br><br>Publication[2] | Purpose / aim | Population:<br><br>Profession (specialist rotation)[1]<br><br>Sample Size[2] | Study Design:<br><br>Comparison group[1]<br><br>pre / post measures[2] | VSP Design:<br><br>Stakeholders Involved<br><br>Pedagogical framework | Desired capabilities | Methodology | Intervention:<br><br>Scenario(s)[1]<br><br>Activities[2]<br><br>Duration[3] | Delivery:<br><br>Software[1]<br><br>Hardware[2] | Student focussed outcome measures | Key Findings | Conclusions/ Implications | Funding / Conflicts of interest |
|---|---|---|---|---|---|---|---|---|---|---|---|---|
| [1] Weston & Zauche (2021)[51]<br><br>[2] USA<br><br>[3] Nurse Educator | To compare the Assessment Technologies Institute (ATI) scores of students who completed their practicum in person versus virtually | [1] 2nd Semester prelicensure baccalaureate nursing students on a paediatric clinical course<br><br>[2] 186 | [1] In-person placement (clinic & simulation) comparator group (n=93)<br><br>[2] Post-test measures | [1] Not specified<br><br>[2] Not mentioned | Take a history<br><br>Perform physical assessment<br><br>Identify problems<br><br>Prioritise interventions<br><br>Integrate foundation knowledge | Quantitative | [1] Physical assessment, sickle cell, cystic fibrosis, infectious respiratory disease, head injury, cardiovascular disease<br><br>[2] Prebrief, i-Human cases, debrief in groups & quizzes<br><br>[3] 5 weeks | [1] Online conferencing (not specified)<br><br>i-Human<br><br>[2] Screen based | Scores on the ATI exam | No difference between the scores on the ATI exam between groups $t(184)=0.700$ ($p=0.485$) | Using the i-Human platform with prebriefing & debriefing, is an effective approach to simulating a pediatric clinical practice | Not stated |
| [1] White et al (2021)[52]<br><br>[2] USA<br><br>[3] Archives of Pathology & Laboratory Medicine | To develop & implement a digital slide–based virtual surgical pathology clinical elective in response to the temporary suspension of in person clinical rotations | [1] Medical students (Pathology rotation)<br><br>[2] 43 | [1] Single group<br><br>[2] Post measures design | [1] Course director (and a needs assessment from student evaluations and rotation data over the proceeding 5 year period)<br><br>[2] Kern's 6 step approach to curriculum development | Summarise the role of a general surgical pathologist.<br><br>List the defining histologic features of several common pathologies.<br><br>Demonstrate how to determine the pathologic stage for an oncologic resection<br><br>Describe how to approach the assessment of a biopsy specimen | Quantitative | [1] Benign & Malignant neopasms. Non-neoplastic, developmental & inflammatory processes<br><br>[2] Reading (e-texts), e-lectures, virtual slides, quizzes, gross dissection videos, student led presentations<br><br>[3] 3 weeks | [1] Zoom PowerPoint Blackboard (LMS) Inversus (Open EdX platform) Internal education web page: hosted the GI pathology module (designed using iSpring Suite 9) & videos. Leica Aperio & Roche iScan (for slide digitisation). Concentriq (for slide delivery). Amazon Web Cloud storage.<br><br>[2] Screen based | Pass / fail assessment.<br><br>Student evaluation survey | All students passed the assessment.<br><br>39.5% survey response rate.<br><br>Learning objectives, patient variety, effective teaching / feedback, the value of technology & the quality of the educational experience were all highly rated | Provided a meaningful clinical experience in a time of online education need.<br><br>Added benefits included increased medical student exposure to pathology as a medical specialty & demonstration of how digital slides can potentially improve standardisation of the pathology clerkship. | None |

| Citation:<br><br>Author (year)¹<br><br>Country²<br><br>Publication³ | Purpose / aim | Population:<br><br>Profession (specialist rotation)¹<br><br>Sample Size² | Study Design:<br><br>Comparison groups¹<br><br>pre / post measures² | VSP Design:<br><br>Stakeholders Involved¹<br><br>Pedagogical Framework² | Desired capabilities | Methodology | Intervention:<br><br>Scenario(s)¹<br><br>Activities²<br><br>Duration³ | Delivery:<br><br>Software¹<br><br>Hardware² | Student focussed outcome measures | Key Findings | Conclusions/ Implications | Funding / Conflicts of interest |
|---|---|---|---|---|---|---|---|---|---|---|---|---|
| [1] Wik et al (2022)[53]<br><br>[2] Canada<br><br>[3] International Journal of Nursing Education Scholarship | To use a community health virtual simulation program to provide clinical placements for undergraduate students | [1] 2nd year Nursing students (community rotation)<br><br>[2] 16 | [1] Single group<br><br>[2] Post-test design | [1] Not specified<br><br>[2] Not stated | Determining factors that impact community health. Gaining insights about diverse health needs, community interventions | Qualitative | [1] Health, social & environmental issues. Infectious disease outbreaks, mental health & cyber bullying<br><br>[2] Observation Applying the community as partner model Windshield surveys Key informant interviews. Planning, implementing & evaluating media campaigns Home assessment Presentations Infographic design Written reflection Quality improvement<br><br>[3] 16 weeks | [1] Sentinel City 3.1 Zoom PowerPoint<br><br>[2] Screen based | All students submitted quality improvement recommendations & 3 of them co-authored a quality improvement assessment with the faculty | Overall, students felt that Sentinel City®3.1 was an adequate program for meeting course objectives. The prescripted design limited the opportunity for critical thinking. | Overall, students who provided feedback considered the platform to be a safe and effective way to teach community and population health nursing concepts and skills. | No funding or conflicts of interest |
| [1] Williams et al (2021)[54]<br><br>[2] USA<br><br>[3] Urology | To design, implement & evaluate learner attitudes of a virtual urologic surgery clinical rotation for medical students. | [1] Senior Medical students (Urology sub-internship)<br><br>[1] 10 | [1] Single group<br><br>[2] Repeated measures design | [1] Not specified<br><br>[2] Aligned with the Urological Association Medical Students Curriculum | Urologic evaluation<br><br>Case presentation<br><br>Anatomy & pathology of common conditions,<br><br>Literature appraisal | Mixed | [1] Benign, oncologic & paediatric urology<br><br>[2] lectures, problem-based learning, reading & videos, discussion board, videoconferences presentations & literature reviews<br><br>[3] 2 weeks | [1] Canvas (LMS)<br><br>BlueJeans (videoconferencing)<br><br>[2] Screen based | Comfort with performing urologic evaluations, confidence in knowledge, identifying conditions & placing consults for urologic issues | Significant ($p<0.05$) increases in: self-perceived knowledge, comfort with performing evaluations, confidence in naming conditions & placing consults | Virtual rotations are scalable & effective at delivering surgical material and can approximate the interpersonal teaching found in clinical learning environments. | Not stated |

| Citation<br><br>Author (year)[1]<br><br>Country[2]<br><br>Publication[3] | Purpose / aim | Population<br><br>Profession (specialist rotation)[1]<br><br>Sample Size[2] | Study Design<br><br>Comparison group[1]<br><br>pre / post measures[2] | VSP Design<br><br>Stakeholders Involved<br><br>Pedagogical framework | Desired capabilities | Methodology | Intervention:<br><br>Scenario(s)[1]<br><br>Activities[2]<br><br>Duration[3] | Delivery:<br><br>Software[1]<br><br>Hardware[2] | Student focussed outcome measures | Key Findings | Conclusions/ Implications | Funding / Conflicts of interest |
|---|---|---|---|---|---|---|---|---|---|---|---|---|
| [1] Zhou et al. (2020)[55]<br><br>[2] China<br><br>[3] Telemedicine and e-health | To observe & analyse the application of Massive Open Online Course (MOOC) & micro video during the COVID-19 epidemic. | [1] Trainee nurses on an Emergency Department (ED) rotation<br><br>[2] 60 | [1] in-person (traditional) placement comparator group (n=30)<br><br>[2] Post-test measures | [1] Nursing Skills Group, including a teaching supervisor, clinical nursing expert & a photographer<br><br>[2] Content was based on the syllabus of the Emergency & Critically Ill Nursing textbook | Theoretical & practical skills with COVID-19 prevention & protection level strategies | Quantitative | [1] Operation of specialised nursing skills involved in the ED<br><br>[2] MOOC & Micro Video course<br><br>[3] 2 weeks | [1] The ED network training platform<br><br>MOOC (based on the textbook)<br><br>[2] Screen based | Theoretical & practical exam scores<br><br>Student evaluation | There was no significant difference in exam scores between groups<br><br>100% survey response rate<br><br>Overall satisfaction, degree of easy understanding, teacher evaluation & learning results group were higher in the experimental group, with statistical significance ($p < 0.05$) | Combined mode of MOOC micro-video can present theoretical and practical courses in a unique way, & is a better alternative when face-to-face and practical courses can no longer be carried out. | None |
