## Supplemental File A6 Mono for "Virtual Simulated Placements in Healthcare Education: A scoping review"

Supplementary File A6: – Papers by Professions

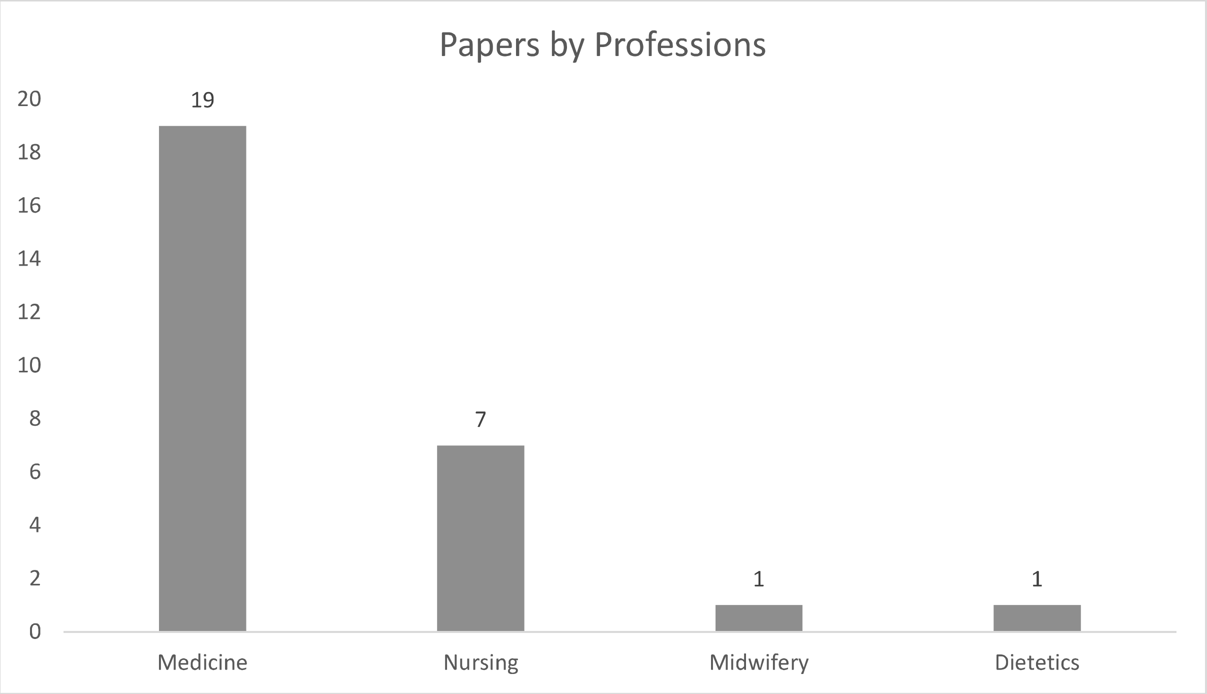


### Breakdown by Medical Specialty

| | |
|---|---|
| Radiology | 4 |
| Emergency | 2 |
| Pathology | 2 |
| Anaesthesiology | 1 |
| GP | 1 |
| Internal Medicine | 2 |
| Neurology | 1 |
| Orthopaedics | 1 |
| Otolaryngology (ENT) | 1 |
| Urology | 1 |
| None | 1 |
| Multiple | 2 |

### Breakdown by Nursing Specialty

| | |
|---|---|
| Paediatrics | 2 |
| Community | 1 |
| Emergency | 1 |
| Psychiatric | 1 |
| Multiple | 1 |
| None | 1 |
