## Supplemental File A7 for "Virtual Simulated Placements in Healthcare Education: A scoping review"

Supplementary File A7: Conceptual Frameworks

| Underpinning Concepts | Type of concept | Papers |
|---|---|---|
| Pedagogy | Student centred learning | Luo et al. (2021)[41] |
| | Andragogy | Bhaysham and Dyer (2020)[29]<br>Creagh et al. (2021)[30] |
| | Problem based learning | Bhaysham and Dyer (2020)[29]<br>Creagh et al. (2021)[30]<br>Kasai et al. (2021)[39] |
| | Experiential learning | Creagh et al. (2021)[30] |
| | Reflective practices | Taylor et al. (2021)[49] |
| | Online learning | Fehl et al. (2022)[33]<br>Villa et al. (2021)[50] |
| Theoretical Frameworks | VSP development | Ganji et al. (2022)[34] |
| | Curriculum development | Nguyen et al. (2023)[43]<br>Redinger and Greene (2021)[45]<br>Samueli et al. (2020)[46]<br>Villa et al. (2021)[50]<br>White et al. (2021)[52] |
| Standards or Existing curricula | International Nursing Association for clinical simulation and learning (USA) | Kubin et al. (2021)[40] |
| | National Standards for Nursing Undergraduates (China) | Luo et al. (2021)[41] |
| | NHS and HCPC Placement standards (UK) | Taylor et al. (2021)[49] |
| | Urological Association of Medical Students Curriculum (USA) | Williams et al. (2021)[54] |
| | Syllabus of the Emergency and Critically Ill Nursing Textbook (China) | Zhou et al. (2020)[55] |
