## Supplemental File A8 for "Virtual Simulated Placements in Healthcare Education: A scoping review"

Supplementary File A8: Bespoke Healthcare Technology

| Type of Resource | Name of Resource | Papers |
|---|---|---|
| Commercial Software (Virtual cases) | Aquifer | Creagh et al. (2021)[30]<br>Durfee et al. (2020)[32] |
| | Body Interact | De Ponti et al. (2020)[31] |
| | vSim | Joung and Kang (2022)[38]<br>Kubin et al. (2021)[40]<br>Luo et al. (2021)[41] |
| | NurseThink vClinical | Kubin et al. (2021)[40] |
| | F.A. Davis Paediatric Interactive Clinical Scenarios for RNs | Kubin et al. (2021)[40] |
| | i-Human | Weston and Zauche (2021)[51] |
| | Sentinel City | Wik et al. (2022)[53] |
| Online modules/training | Cleveland Clinic Paediatric Radiology Modules | Gomez et al. (2020)[35] |
| | Virtual Healthcare Experience | Kubin et al. (2021)[40] |
| | National Prescribing Module | Smith and Jones (2023)[47] |
| | Online MedEd CaseX | Redinger and Greene (2021)[45] |
| | Emergency Medicine Reviews and Perspectives | Redinger and Greene (2021)[45] |
| | Diet-COMMS | Taylor et al. (2021)[49] |
| | IDHEAL Modules | Villa et al. (2021)[50] |
| | Foundations of EM | |
| | Scientific Foundations of Medicine (SFM) histology and pathobiology | White et al. (2021)[52] |
| Custom built sites/apps | Gastrointestinal (GI) Pathology | |
| | Internal trainee education page | White et al. (2021)[52] |
| | Google Sites | Kubin et al. (2021)[40] |
| | Articulate.com | Rahm et al. (2021)[44] |
| | 360 images | Taylor et al. (2021)[49] |
| | The ED Network Training Platform | Zhou et al. (2021)[55] |
| Online resources | Pacsbin<br>TeamRads.com<br>LearningRadiology.com<br>CTisUs.com<br>High Value Practice Academic Alliance Ordering Wisely E-Lectures | Gomez et al. (2020)[35] |
| | One Night in the ED | Gomez et al. (2020)[35]<br>Redinger and Greene (2021)[45] |
| | Sublux Radiology | Redinger and Greene (2021)[45] |
| | John Hopkins Surgical Pathology (login required) | White et al. (2021)[52] |
| Whole slide image viewers | Concentriq | |
| | CaseViewer<br>Aperio ImageScope | Samueli et al. (2020)[46] |
